# Nanopore 16S rRNA sequencing reveals alterations in nasopharyngeal microbiome and enrichment of *Mycobacterium* and *Mycoplasma* in patients with COVID 19

**DOI:** 10.1101/2021.11.10.21266147

**Authors:** Soumendu Mahapatra, Rasmita Mishra, Punit Prasad, Krushna Chandra Murmu, Shifu Aggarwal, Manisha Sethi, Priyanka Mohapatra, Arup Ghosh, Rina Yadav, Hiren Dodia, Shamima Azma Ansari, Saikat De, Deepak Singh, Amol Suryawanshi, Rupesh Dash, Shantibhushan Senapati, Tushar K. Beuria, Soma Chattopadhyay, Gulam Hussain Syed, Rajeeb Swain, Sunil K. Raghav, Ajay Parida

**Author notes:** Correspondence: **Ajay Parida, Ph.D.**, Institute of Life Sciences, Nalco Square, Chandrasekharpur, Bhubaneswar, Odisha – 751023, **Punit Prasad, Ph.D.**, Institute of Life Sciences, Nalco Square, Chandrasekharpur, Bhubaneswar, Odisha – 751023. These authors contributed equally to this work (Soumendu Mahapatra, Rasmita Mishra, Punit Prasad).

## Abstract

The coronavirus disease 2019 (COVID-19) pandemic caused by severe acute respiratory syndrome corona virus 2 (SARS-CoV-2) is a major global health concern. This virus infects the upper respiratory tract and causes pneumonia-like symptoms. So far, few studies have shown that respiratory infections alter nasopharyngeal (NP) microbiome diversity and enrich opportunistic pathogens. In this study, we have sequenced the 16S rRNA variable regions, V1 through V9, extracted from NP samples of control and COVID-19 (symptomatic and asymptomatic) participants using the Oxford Nanopore™ technology. Comprehensive bioinformatics analysis investigating the alpha/beta diversities, non-metric multidimensional scaling, correlation studies, canonical correspondence analysis, linear discriminate analysis, and dysbiosis index analysis revealed control and COVID-19-specific NP microbiomes. We observed significant dysbiosis in COVID-19 NP microbiome with abundance of opportunistic pathogens such as *Cutibacterium, Corynebacterium, Oerskovia*, and *Cellulomonas* in asymptomatic patients, and of *Streptomyces* and *Mycobacteriaceae* family in symptomatic patients. Furthermore, we observed sharp rise in enrichment of opportunistic pathogens in symptomatic patients, with abundance of *Mycobacteria* and *Mycoplasma*, which strongly correlated with the occurrences of chest pain and fever. Our findings contribute novel insights regarding emergence of opportunistic pathogens in COVID-19 patients and their relationship with symptoms, suggesting their potential role in coinfections.

## Introduction

The coronavirus disease 2019 (COVID-19) pandemic, a global health threat, is caused by severe acute respiratory syndrome coronavirus 2 (SARS-CoV-2). The symptoms range from fever, throat pain, loss of taste and smell to severe congestion in the chest, drop in oxygen levels, pneumonia, and acute respiratory distress syndrome (1). Furthermore, a significant population worldwide remains asymptomatic, which is considered spreaders of the infection (2). The virus enters the host via the upper respiratory tract (URT) where the spike protein binds to the angiotensin I converting enzyme 2 (ACE2) receptor, an essential step in invading host cells to cause progressive disease (3, 4). Random mutations in the SARS-CoV2 spike protein and receptor-binding domain promote efficient invasion and enhance pathogenicity (5).

The nasopharyngeal tract is inhabited by a large number of microbial communities which maintain normal homeostasis (6). Studies have revealed association between microbial communities that influence viral infections of the lung, such as chronic rhinosinusitis, asthma, pneumonia, and cystic fibrosis in the URT (7, 8). URT microbiome dysbiosis may also enhance the opportunistic pathogen population and promote coinfection in the host (9, 10). Reports have shown that nasopharyngeal (NP) swabs in viral transport media can be used to investigate the NP microbial composition in patients with COVID-19 (11, 12). Recent studies have revealed overall compositional changes in the NP microbiota and promotion of opportunistic pathogens such as *Rothia* and *Veillonella* in COVID-19 patients with shortness of breath (11, 13, 14). The secondary infection in patients with COVID-19 is associated with abundance of opportunistic pathogens such as *Moraxella, Corynebacterium, Haemophilus, Stenotrophomonas, Acinetobacter, Fusobacterium periodonticum*, and *Pseudomonas aeruginosa* (15-18). Studies on functional pathways of the NP metagenomics have revealed that the abundance of NP commensal bacteria such as *Gemella morbillorum, Gemella haemolysans*, and *Leptotrichia hofstadii* was reduced in the respiratory tract of COVID-19 patients, indicating the role of distinct functional metabolic pathways in this infection (19, 20). Little is known about the crosstalk between SARS-CoV-2 viral infection and NP microbiota. Moreover, systematic data connecting COVID-19-associated symptoms with microbial composition is lacking. The absence of an animal model makes it difficult to test and validate the role of NP microbiota in SARS-CoV-2 infection. Studies so far have shown differences in the abundance of different opportunistic pathogens in the NP microbiota of patients, which is one of the bottlenecks in this area of research. Hence more studies on the NP microbiome are required for understanding its role in symptomatic and asymptomatic COVID-19 patients and its relation with symptom severity.

In this study, we have investigated the alterations in the NP microbial ecosystem of patients with active COVID-19 (n = 46) and compared them with that of healthy individuals (n = 12). We have used the 16S metagenome approach and long-read sequencing (V1−V9) with the Nanopore sequencing method to elucidate the reduction in microbial diversity in patients with COVID-19. The composition of the NP microbiota changed significantly between symptomatic and asymptomatic patients, resulting in enrichment of opportunistic pathogens Interestingly, we found abundance of *Mycoplasma* and *Mycobacterium* at the genus level, which strongly correlated with chest pain and fever in the symptomatic patients.

## Materials and Methods

### Ethical approval

Ethical permission for nasopharyngeal microbiome study and the biorepository was obtained from the Institutional Ethical Committee (IEC)/Institutional Review Board (IRB) of the Institute of Life Sciences [(102/HEC/2020) and (100/HEC/2020)]. Approval was also obtained from the Institutional Biosafety Committee (IBSC) (V-122-MISC/2007-08/01/2/2.1) for this study and the biorepository (V-122-MISC/2007-08/01) and from the Review Committee on Genetic Manipulations (RCGM) under Department of Biotechnology, Ministry of Science and Technology.

### Sample collection and reverse transcription-polymerase chain reaction (RT-PCR)

In total, 60 NP samples were collected for 16S rDNA amplicon sequencing from the Institute of Life Science (ILS) COVID-19 sample biorepository unit. The COVID-19-positive samples (n = 47) were confirmed by amplifying the genes encoding SARS-CoV-2 nucleocapsid, spike, and ORF1ab/RdRP using either TaqPath™ COVID-19 combo kit (Invitrogen, A47814) or Meril COVID-19 one-step RT-PCR kit (Meril Diagnostics, NCVPCR-02). All samples were collected in the hospital setup prior to the medication. These COVID-19-positive patients were not treated with antibiotics as the patients were not aware of their COVID-19 testing results. The COVID-19 patients were grouped as symptomatic (n = 22) or asymptomatic (n = 25) based on their clinical data. The control samples (n = 13) were negative for SARS-CoV-2 virus RNA and none of the subjects from whom the samples were obtained had any flu-like symptoms. All samples were collected in viral transport media (VTM) and stored at –80°C until DNA isolation.

### DNA extraction and PCR amplification

DNA was isolated using the PureLink™ microbiome DNA purification kit (Invitrogen, A29790) according to the manufacturer’s protocol and eluted in 40 μl elution buffer. The quality and quantity of DNA were determined using the Multiskan™GO spectrophotometer (Thermo Scientific). 16S rDNA amplification, library preparation, and sequencing: V1-V9 variable regions of the 16S rRNA gene were amplified using 130-F (5′-GGCGGATCCAAGGAGGTGTTCCAGCCGC-3′) and 139-R (5′-GGCCTCGAGAGAGTTTGATCCTGGCTCAGG-3′) primers. PCR (50 μl) was set up using total DNA (10 ng) isolated from NP samples, primers (5 nM), and NEB Q5® High-Fidelity 2 X master mix (NEB, M0492L) per the manufacturer’s protocol. The amplicons (∼1.6 kb) were analyzed on 0.8% agarose gel and cleaned using DNA Clean and Concentrator-25 kit (Zymo Research, D4034). The PCR products were quantified using a Qubit 4 fluorometer (Thermo Scientific) using the Qubit® dsDNA BR assay kit (Thermo Scientific, Q32853). Amplicon libraries were generated following the Oxford Nanopore 1D library preparation protocol using the PCR barcoding (96) genomic DNA kit (Oxford Nanopore™, SQK-LSK109). Equimolar amounts of amplicon libraries were pooled and sequenced using the MinION OXFORD NANOPORE™ device at the ILS DNA sequencing facility.

### Microbiome data processing

RAW fast5 files were generated using the MinKNOW™ tool for individual samples. Base calling was performed using the Guppy base-caller and fastq files were generated. FastQC of each sample was performed using the Babraham fastqc suite (https://www.bioinformatics.babraham.ac.uk/projects/fastqc/), followed by trimming of low quality reads using nanoflit. Operational taxonomic units (OTUs) were generated using Kraken2 (https://ccb.jhu.edu/software/kraken2/index.shtml) (21) and the unclassified reads were filtered for downstream analysis using the ‘phyloseq’ ‘R” package to generate combined OTUs for all the samples and metadata (Supplemental Table 1). Read counts for mitochondria and chloroplast were discarded. Normalization and differential OTU abundance were determined between control, and symptomatic and asymptomatic subjects using the DESeq2 function (cutoff of p-value ≤ 0.05). The accession ID in NCBI is PRJNA774098.

### In-depth microbiome data analysis

#### Diversity analysis

Alpha diversity was assessed using the Shannon diversity index and Simpson Diversity index. Statistical significance was estimated using the Wilcoxon rank sum test. The beta diversity significance among groups was examined with PERMANOVA (p-value 0.001). Ordination analysis was performed by PCoA, NMDS and CCA. R packages used are ‘microbiome’,’Vegan’,’ade4’,ggpubr for analysis and ‘ggplot2’ for visualization.

#### Dysbiosis index

Microbiome dysbiosis in each sample was calculated based on Bray-Curtis distances. All samples were subjected to PCoA using Bray-Curtis distances. Next, the centroid (median) of the control subjects was calculated along PCoA axes. The dysbiosis score for each sample was calculated as a Euclidian distance between its position in the PCoA space and control centroid 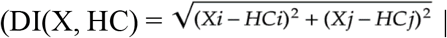 (DI: Dysbiosis Index, X: Samples, HC: Control Centroid). Their significance was assessed using Wilcoxon and Kruskal-Wallis test (22).

#### Sample correlation

Correlation matrix between samples and OTUs for each taxonomic level (phylum, order, family, and genus) from differential OTUs was obtained using Spearman’s correlation method and it was visualized as a heat map. Correlation coefficients for each sample correlation pair and each classification level and density plot were plotted with mean and median. The Kolmogorov test (KS) was used to determine the significance in sample groups (control, asymptomatic, and symptomatic).

#### Linear discriminant analysis (LDA) effect size (LEfSe) analysis

The LEfSe was calculated using the online Galaxy web application with the Huttenhower lab’s tool (https://huttenhower.sph.harvard.edu/galaxy/). LDA effect size was calculated using the Kruskal-Wallis sum rank test (alpha = 0.05) and it detected differential abundant features at genus and species level within three sample groups. The taxonomic-level significance was then tested using the pairwise Wilcoxon rank-sum tests (alpha = 0.05). Finally, the effect size of each differentially abundant feature was estimated using LDA. One-against-all sample groups were compared and a linear discriminant analysis score greater than 3.6 was set as the threshold; all-against-all sample groups were compared and a linear discriminant analysis score greater than 2.0 was set as the threshold. Cladogram was used for identification of taxa at different levels of the taxonomic hierarchy between sample groups (LDA score > 2).

#### Network analysis

Network was constructed using weighted correlation network analysis or weighted gene co-expression network analysis (WGCNA). Briefly, pairwise Spearman correlation between OTUs (which was generated from LefSe analysis) was calculated using the WGCNA function. Network metrics such as betweenness, closeness, Eigen centrality, and PageRank centrality of the resulting network were calculated and visualized using ‘Gephi’, (https://gephi.org/) (23).

## Results

### Study design and subject attributes

The role of the microbiome in viral infections is an emerging field. We collected NP samples from COVID-19 patients between 11th May 2020 and 10th October 2020 to study alterations in the NP microbiome. The schematic representation of the nasal microbiome study with 16S rDNA amplicon sequencing is shown in Figure 1A. In total, 60 NP samples subjects (infected, n = 47 and control, n = 13 subjects, positive and negative for SARS-CoV2 RT-qPCR test respectively) were obtained from the Institute of Life Sciences biorepository. Out of 47 SARS-CoV-2-positive subjects, 25 were asymptomatic and 22 were symptomatic with mild symptoms (Figure 1B). In total, 179,59,691 reads were generated. Two samples with low read counts (1 from control and other from symptomatic category) were excluded and the final study was performed with 58 subjects, including the control (C) [n = 12 (21%)], asymptomatic [IA, infected asymptomatic; n = 25 (43%)], and symptomatic [IS, infected symptomatic; n = 21 (36%)]. The details of the participants considered for this study are described in Table 1. Differential OTUs (n = 795, p ≤ 0.05) were obtained from a total of 3482 OTUs using the deseq2 function by comparing with control NP subjects. For downstream analysis differential, 795 OTUs were considered. We used the t-distributed stochastic neighbor embedding (t-SNE) dimension reduction method to obtain the overall distribution of NP samples with 795 OTUs (Figure 1C). We found that the control and SARS-CoV-2-infected subjects showed distinct segregation of OTUs in the NP microbiome, while asymptomatic and symptomatic subjects showed modest separation. This indicated that the abundance of 795 differential OTUs potentially determines the compositional distribution patterns.

**Table 1:** Details of samples included in this study.

|  | **Control (n=12)** | **Asymptomatic (n=25)** | **Symptomatic (n=21)** |
| --- | --- | --- | --- |
| **Sex** |  |  |  |
| Male | 5 (41.66%) | 18 (72%) | 19 (90.47%) |
| Female | 7 (58.33%) | 7 (28%) | 2 (9.52%) |
| Age (years) | 31 (median) | 26 (median) | 32 (median) |
| **Symptoms** |  |  |  |
| Dry Cough | NA | NA | 7 (33.3%) |
| Fever | NA | NA | 17 (80.95%) |
| Tiredness | NA | NA | 8 (38.09%) |
| Sore throat | NA | NA | 11(52.38%) |
| Body pain | NA | NA | 13(61.9%) |
| Chest pain | NA | NA | 9(42.85%) |
| Fever + Body pain | NA | NA | 4(19.04%) |
| Fever + multiple symptoms* | NA | NA | 16(76.19%) |
| Fever + chest pain | NA | NA | 8(38.09%) |
| Loss of smell/taste + multiple symptoms* without fever | NA | NA | 2(9.52%) |

\*Multiple symptoms refer to having more than one symptoms from symptoms list.

**Figure 1:**
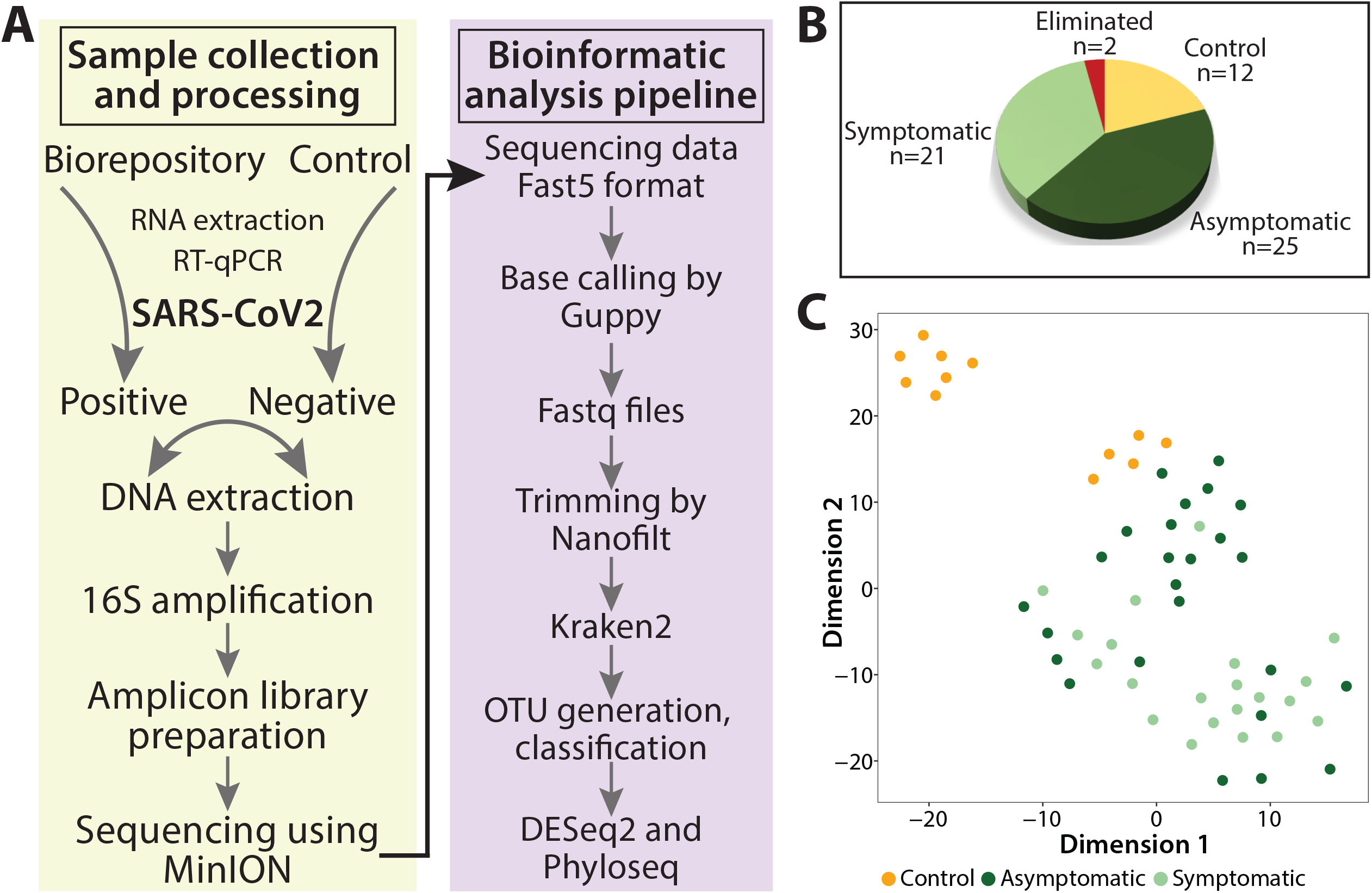
Schema of nasopharyngeal sample processing, 16S sequencing, and OTU-based sample distribution. (A) Flow chart showing nasopharyngeal sample processing for DNA extraction, amplicon library preparation, Oxford Nanopore™ sequencing, and bioinformatics analysis pipeline. (B) Pie chart showing nasopharyngeal samples (controls, symptomatic, and asymptomatic) used in this study. (C) t-SNE plot showing the OTU-based sample distribution and ordination points for control, symptomatic, and asymptomatic samples.

### NP microbiome diversity was significantly altered in COVID-19 patients

Distinct distribution of OTUs from control and infected patients prompted us to compare the evenness and richness of bacterial community compositions using Shannon and Simpson alpha indices. The Shannon and Simpson alpha microbial diversity indices between control and SARS-CoV-2-infected participants differed significantly (p-value ≤ 0.05) in pairwise Wilcoxon rank test (Shannon p-value =3.0 × 10^−4^ and Simpson p-value=3.3 × 10^−3^) (Figure 2A, B). Although the alpha diversity indices for samples from symptomatic and asymptomatic patients compared to control subjects were found to be significantly reduced, no difference was observed between symptomatic and asymptomatic samples (Figure 2C, D). Furthermore, we used a linear regression model to establish the association between total OTU read counts for each sample and Shannon/Simpson alpha diversity indices. We found negative correlation for both Shannon (IA - R = −0.35, R^2^ = 0.44, p = 0.083; IS - R = −0.54, R^2^ = 0.48, p = 0.012) and Simpson (IA - R = −0.58, R^2^ = 0.68, p = 0.0028; IS - R = −0.77, R^2^ = 0.63, p = 7.7 × 10^−5^) alpha diversity indices with 95% confidence intervals with total OTU counts (Figure 2E, F). To further understand the microbial composition dissimilarity within the samples, we analyzed beta diversity using principal coordinate analysis (PCoA) and applied both unweighted (microbial richness) and weighted (microbial richness and abundance) unifrac distance methods. The first two components of PCoA showed 60.3% and 80.1% variance for the unweighted and weighted unifrac method. The overall difference in microbial population showed two different clusters of control and SARS-CoV-2-infected patients (IA and IS) in the unifrac weighted method, while the unifrac unweighted method showed more clear segregation between symptomatic and asymptomatic samples (Figure 2G). We assessed the significance of beta diversity to calculate unifrac distance matrix (PERMANOVA test with 999 permutations) for both unweighted and weighted methods and found that the three sample groups (C, IA, and IS) differed significantly (P = 0.001) with 18% variance explained (R^2^ = 0.18842).

**Figure 2:**
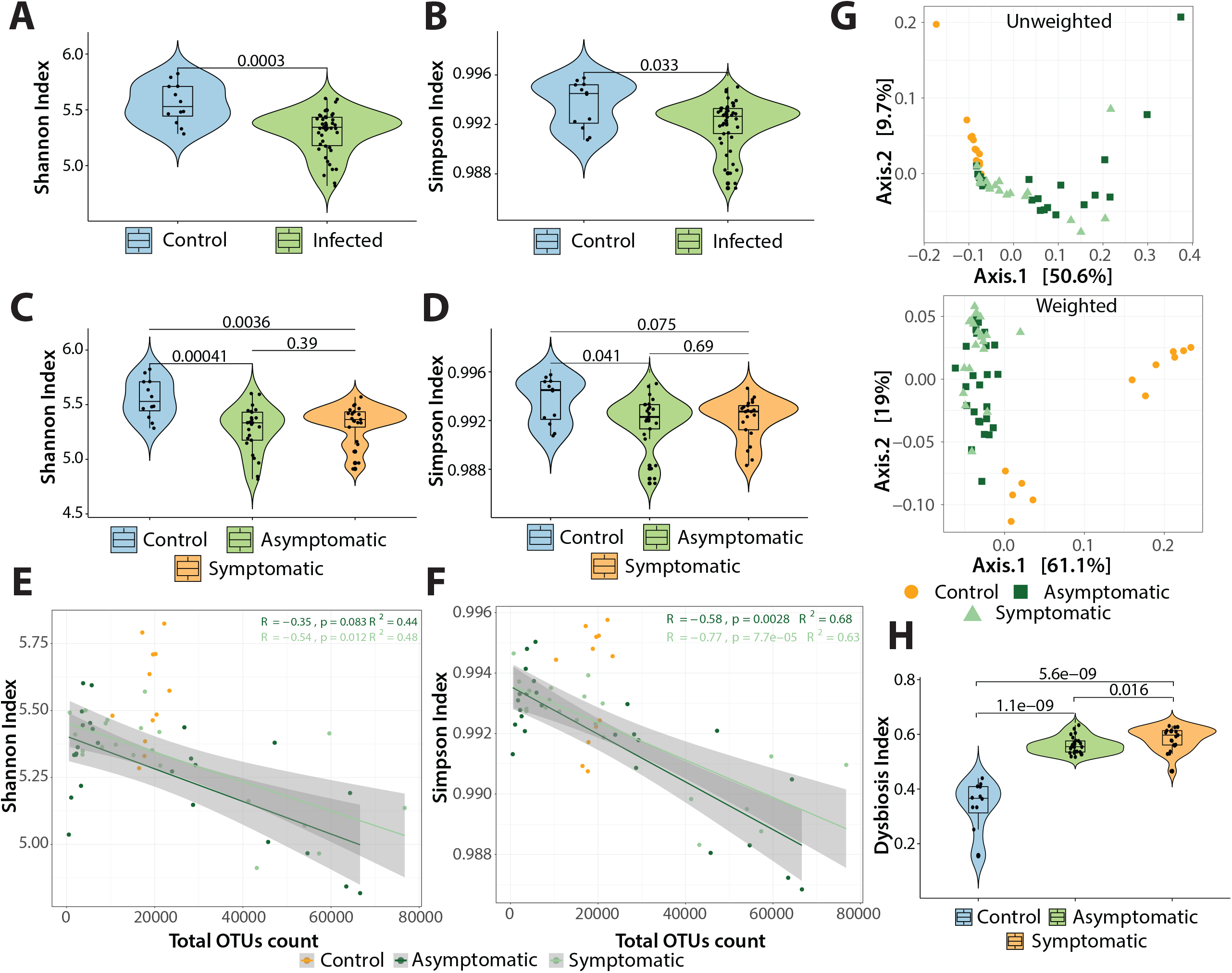
Alpha/beta diversities and dysbiosis index in COVID-19-positive and negative nasopharyngeal sample. (A-B) Alpha diversity index (Shannon/Simpson) between control and COVID-19-infected samples (pairwise Wilcoxon rank-sum test p = ≤0.05). (C-D) Same analysis as above where the COVID-19-infected samples are classified as asymptomatic and symptomatic compared to the control group. (E-F) Linear regression model showing the association between total OTU count and Shannon/Simpson diversity index for each sample; the shaded grey region represents 95% confidence intervals of two groups, symptomatic and asymptomatic, with correlation (Spearman) regression line [Shannon: R = −0.35 (asymptomatic), R = −0.54 (symptomatic) and Simpson: R = −0.58 (asymptomatic), R = −0.77 (symptomatic)]. (G) Principal coordinate analysis (PCoA) showing beta diversity in asymptomatic, symptomatic, and control sample groups based on unifrac (weighted/unweighted) distance (p = 0.001, PERMANOVA). (H) Violin plot showing dysbiosis indexes of samples from control, asymptomatic, and symptomatic participants (pairwise Wilcoxon rank-sum test p = ≤0.05).

### NP microbiome dysbiosis in COVID-19 patients

Alterations in the microbial diversity prompted us to determine microbial dysbiosis index (DI) (alterations in the microbial community) across the three groups (C, IA, and IS). We performed PCoA using the Bray Curtis distance matrix and found that NP microbiota was significantly altered (p = 0.001) with 61% variation in distances explained (R^2^ = 0.6136) assessed by ADONIS test. Next, we calculated the Euclidean distance from the centroid for samples from control (median = 0.3404), asymptomatic (median = 0.1881) and symptomatic (median = 0.1511) individuals and calculated the DI (Supplementary figure 1B). The overall observed DI was significant (Kruskal-Wallis test, p = 1.317E-07) across all the groups. Pairwise comparison showed significant dysbiosis between control vs symptomatic (p = 5.6E−09) and control vs. asymptomatic (p = 1.1E−09) groups; however, dysbiosis between asymptomatic and symptomatic (p = 0.016) pair was not highly significant (Figure 2H). We also observed highly significant dysbiosis (p = 2.2E-12) between the control and infected group (Supplemental Figure 1A, 1C). This showed that compared to that in the control subjects, the NP microbial community is severely altered in both symptomatic and asymptomatic COVID-19 patients.

### Distinct microbial composition and abundance at phylum and family levels in patients suffering from SARS-CoV2 infection

The alpha and beta diversities, and DI showed that the NP microbiome was significantly altered in COVID-19 patients. Next, we aimed to identify the microbial communities that were altered at the phylum and family levels in three sample groups. We found 795 differential OTUs, out of which, 12 phyla, 65 orders, 126 families, and 240 genera were present in all three groups (C, IA, and IS) (Supplemental Table 1). The 12 phyla and their significance is shown in Table 2. The most significant bacteria in phylum level were Actinobacteria (p = 9.96E-07) and Proteobacteria (p = 9.61E-07), including 9 other phyla assessed using the Kruskal-Wallis test. The abundance of phyla Firmicutes (p = 4.65E-02) and Actinobacteria (p = 9.96E-07) were significantly higher in the SARS-CoV-2-infected groups (symptomatic and asymptomatic). In contrast, Bacteroidetes (p = 1.48E-06) and Proteobacteria (p = 6.56E-07) were highly abundant in the control group (non-infected) (Supplemental Figure 2A). Furthermore, we analyzed relative abundance of top 30 families and found enrichment of *Mycobacteriaceae, Propionibaceriaceae*, and *Streptomycetaceae* (Supplemental Figure 2B). These families contain opportunistic pathogens in both symptomatic and asymptomatic COVID-19 patients, while these families are absent in control subjects. Top families and their significance is shown in Table 3.

**Table 2:** Phylum based on relative abundance and their respective values.

| **Phylum** | **Mean** | **1st Quartile** | **Median** | **3rd Quartile** | **p_value** | **BH_FDR** |
| --- | --- | --- | --- | --- | --- | --- |
| Proteobacteria | 1.51E-01 | 3.79E-02 | 4.53E-02 | 7.29E-02 | 6.56E-07 | 6.56E-07 |
| Fusobacteria | 2.25E-03 | 1.31E-03 | 1.72E-03 | 2.71E-03 | 9.00E-07 | 9.00E-07 |
| Actinobacteria | 6.83E-01 | 7.09E-01 | 7.62E-01 | 7.99E-01 | 9.96E-07 | 9.96E-07 |
| Bacteroidetes | 8.77E-03 | 5.71E-03 | 6.66E-03 | 1.00E-02 | 1.48E-06 | 1.48E-06 |
| Tenericutes | 9.37E-04 | 1.35E-04 | 5.60E-04 | 1.21E-03 | 2.21E-06 | 2.21E-06 |
| Chloroflexi | 2.25E-03 | 6.47E-04 | 9.84E-04 | 1.46E-03 | 6.54E-06 | 6.54E-06 |
| Chlamydiae | 5.93E-04 | 2.82E-04 | 4.98E-04 | 7.76E-04 | 7.27E-06 | 7.27E-06 |
| Fibrobacteres | 5.65E-04 | 2.86E-04 | 4.05E-04 | 7.10E-04 | 4.77E-05 | 4.77E-05 |
| Thermodesulfobacteria | 2.62E-02 | 2.09E-02 | 2.84E-02 | 3.24E-02 | 8.13E-04 | 8.13E-04 |
| Aquificae | 2.31E-04 | 1.19E-04 | 2.30E-04 | 2.93E-04 | 1.59E-03 | 1.59E-03 |
| Firmicutes | 1.23E-01 | 1.19E-01 | 1.28E-01 | 1.44E-01 | 4.65E-02 | 4.65E-02 |
| Chlorobi | 1.02E-03 | 6.81E-04 | 9.32E-04 | 1.27E-03 | 8.58E-01 | 8.58E-01 |

**Table 3:** Top 30 family based on relative abundance and their respective values.

| Family | Mean | 1st quartile | Median | 3rd quartile | p_value | BH_FDR |
| --- | --- | --- | --- | --- | --- | --- |
| *Acetobacteraceae* | 5.08E-02 | 5.00E-02 | 5.12E-02 | 5.28E-02 | 9.14E-07 | 9.14E-07 |
| *Actinomycetaceae* | 2.23E-02 | 2.10E-02 | 2.18E-02 | 2.32E-02 | 8.18E-03 | 8.18E-03 |
| *Aeromonadaceae* | 2.96E-02 | 2.54E-02 | 3.07E-02 | 3.38E-02 | 8.18E-03 | 8.18E-03 |
| *Alcaligenaceae* | 8.14E-02 | 8.50E-02 | 8.71E-02 | 9.43E-02 | 1.41E-06 | 1.41E-06 |
| *Bifidobacteriaceae* | 7.69E-02 | 4.59E-02 | 6.52E-02 | 1.04E-01 | 2.99E-03 | 2.99E-03 |
| *Brevibacteriaceae* | 4.37E-02 | 2.72E-02 | 3.82E-02 | 5.98E-02 | 6.35E-04 | 6.35E-04 |
| *Cellulomonadaceae* | 6.75E-02 | 4.60E-02 | 6.29E-02 | 8.52E-02 | 8.64E-05 | 8.64E-05 |
| *Chromobacteriaceae* | 2.30E-02 | 2.27E-02 | 2.30E-02 | 2.32E-02 | 9.37E-07 | 9.37E-07 |
| *Corynebacteriaceae* | 5.49E-02 | 4.18E-02 | 5.05E-02 | 6.52E-02 | 2.36E-05 | 2.36E-05 |
| *Enterobacteriaceae* | 1.26E-01 | 3.61E-02 | 1.65E-01 | 2.17E-01 | 1.73E-06 | 1.73E-06 |
| *Erwiniaceae* | 3.19E-02 | 2.97E-02 | 3.10E-02 | 3.46E-02 | 2.01E-06 | 2.01E-06 |
| *Erysipelotrichaceae* | 2.26E-02 | 2.21E-02 | 2.26E-02 | 2.30E-02 | 1.69E-06 | 1.69E-06 |
| *Eubacteriaceae* | 2.99E-02 | 2.58E-02 | 2.84E-02 | 3.28E-02 | 1.85E-06 | 1.85E-06 |
| *Lactobacillaceae* | 2.12E-02 | 2.05E-02 | 2.10E-02 | 2.18E-02 | 5.39E-03 | 5.39E-03 |
| *Micrococcaceae* | 3.52E-02 | 2.65E-02 | 3.37E-02 | 4.36E-02 | 3.49E-03 | 3.49E-03 |
| *Micromonosporaceae* | 2.41E-02 | 2.25E-02 | 2.43E-02 | 2.53E-02 | 2.65E-06 | 2.65E-06 |
| *Morganellaceae* | 5.01E-02 | 4.51E-02 | 5.30E-02 | 5.93E-02 | 1.90E-05 | 1.90E-05 |
| *Mycobacteriaceae* | 2.19E-01 | 1.85E-01 | 2.42E-01 | 2.69E-01 | 7.93E-07 | 7.93E-07 |
| *Neisseriaceae* | 5.24E-02 | 5.08E-02 | 5.23E-02 | 5.43E-02 | 1.29E-06 | 1.29E-06 |
| *Nocardiaceae* | 3.15E-02 | 2.46E-02 | 3.41E-02 | 3.63E-02 | 8.90E-07 | 8.90E-07 |
| *Pasteurellaceae* | 4.73E-02 | 3.34E-02 | 5.14E-02 | 5.75E-02 | 5.89E-05 | 5.89E-05 |
| *Pectobacteriaceae* | 2.17E-02 | 2.05E-02 | 2.15E-02 | 2.27E-02 | 2.09E-06 | 2.09E-06 |
| *Peptostreptococcaceae* | 3.01E-02 | 2.56E-02 | 2.82E-02 | 3.23E-02 | 1.89E-06 | 1.89E-06 |
| *Propionibacteriaceae* | 1.17E-01 | 9.00E-02 | 1.20E-01 | 1.33E-01 | 8.90E-07 | 8.90E-07 |
| *Pseudonocardiaceae* | 2.94E-02 | 2.70E-02 | 2.98E-02 | 3.27E-02 | 1.44E-06 | 1.44E-06 |

### Taxonomic classifications based on OTU abundance showed sample group segregation at the genus level

To further our understanding regarding the 795 differentially abundant OTUs, we used the NMDS approach at phylum, order, family, and genus levels for C, IA, and IS sample groups using the Bray-Curtis distance matrix. Statistical significance using ANOSIM for phylum (R = 0.262, p = 1.7E-03), order (R = 0.322, p = 3E-04), family (R = 0.3461, p = 3E-04), and genus (R = 0.3507, p = 3E-04) showed gradual increase in R-value for genus. This indicated that as we go lower in the taxonomic classification, the variance in the OTUs provides better sample segregation. The differential OTUs present at the genus level in three sample groups have high level of dissimilarity (35%) with R = 0.3507 and show clear sample segregation (Figure 3A). To further validate the NMDS findings and identify the NP OTU differences between C, IA, and IS sample groups, we used sample correlation (Spearman matrix) (Supplemental Figure 3A-D). The sample correlation matrix clearly showed distinction among C, IA, and IS with respect to taxonomic classification (Figure 3B). To further reconcile the distinct sample segregation at higher to lower taxonomic level based on OTU abundances, we plotted density histogram of correlation coefficient values (obtained in sample correlation). The mean and median value of each density plot revealed lack of difference between the C, IA and IS groups at the phylum level. Furthermore, subtle differences were observed at the order and family level. However, at the genus level, we found comprehensible differences between C (mean = 7.95E-01; median = 6.39E-01), IA (mean = 5.65E-01; median = 8.33E-01) and IS (mean = 6.51E-01; median = 7.01E-01) (Table 4) (Figure 3B). To evaluate the statistical significance of densities based on sample segregation, we calculated cumulative distribution distance (D) and significance between C, IA, and IS groups using the Kolmogorov–Smirnov (KS) test for each taxonomic rank (Table 5). We observed that compared to that at other taxonomic levels, all the comparisons were highly significant at the genus level. Based on the ‘D’ value comparison the samples were well distributed in C vs. IA (D = 5.94E-01; p-value < 2.2E-16), C vs. IS (D = 5.06E-01; p = 3.308E-14), and IA vs. IS (D = 2.28E-01; p = < 2.2E-16) at the genus level. The overall sample distribution differences were highly enriched at the genus level than between these three groups. Although the ‘D’ value between IA and IS groups was less but the distribution pattern showed significant differences between them. The above observance enables us to consider the genus level OTUs (n = 240) for downstream analysis.

**Table 4:** Mean and median value of density plots.

| Taxonomic rank | Statistic value | Control | Asymptomatic | Symptomatic |
| --- | --- | --- | --- | --- |
| Phylum | Mean | 8.96E-01 | 8.68E-01 | 8.90E-01 |
|  | Median | 9.30E-01 | 8.95E-01 | 9.16E-01 |
| Order | Mean | 8.58E-01 | 7.70E-01 | 8.36E-01 |
|  | Median | 8.78E-01 | 8.21E-01 | 8.93E-01 |
| Family | Mean | 8.22E-01 | 6.86E-01 | 7.60E-01 |
|  | Median | 8.63E-01 | 7.46E-01 | 8.06E-01 |
| Genus | Mean | 7.95E-01 | 5.65E-01 | 6.51E-01 |
|  | Median | 6.39E-01 | 8.33E-01 | 7.01E-01 |

**Table 5:** Result of Kolmogorov–Smirnov (KS) test between the densities of each taxonomic rank.

| **Taxonomic rank** | **Control vs Asymptomatic** | **Control vs Symptomatic** | **Asymptomatic vs Symptomatic** |
| --- | --- | --- | --- |
| **Phylum** | D = 1.88e-01 | D = 1.07e-01 | D = 1.01e-01 |
|  | p-value = 3.03e-02 | p-value = 0.4834 | p-value = 9.78e-04 |
| **Order** | D = 3.18e-01 | D = 9.97e-02 | D = 2.30e-01 |
|  | p-value = 1.237e-05 | p-value = 0.5706 | p-value < 2.2e-16 |
| **Family** | D = 4.14e-01 | D = 2.93e-01 | D = 2.25e-01 |
|  | p-value = 2.706e-09 | p-value = 4.75e-05 | p-value < 2.2e-16 |
| **Genus** | D = 5.94e-01 | D = 5.06e-01 | D = 2.28e-01 |
|  | p-value < 2.2e-16 | p-value = 3.308e-14 | p-value < 2.2e-16 |

**Figure 3:**
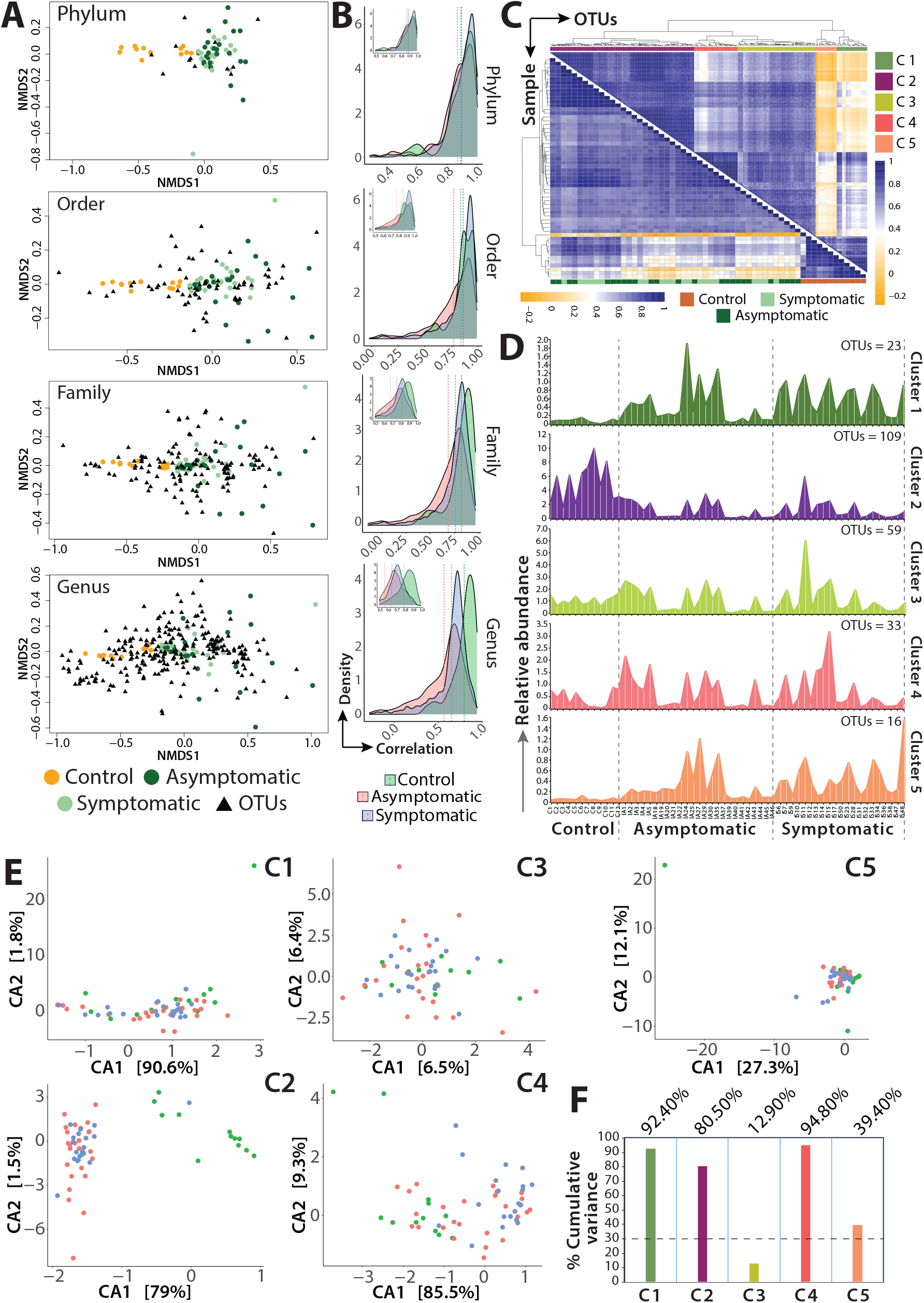
Taxonomic classification of bacterial communities using non-metric multidimensional scaling (NMDS), correlation, and canonical correspondence analysis (CCA). (A) NMDS ordination of Bray-Curtis distance matrix based on all samples and bacterial communities of each taxonomy level (phylum, order, family, and genus) (ANOSIM p = <0.05). (B) The density plot representing the Spearman correlation coefficient at each taxonomy level (phylum, order, family, and genus); dotted line indicates the mean value of each sample group (Kolmogorov–Smirnov (KS) Test p ≤ 0.05). (C) Heat map of Spearman correlation for genus level with sample correlation (lower) and OTU correlation (upper). Five clusters (C1, C2, C3, C4, and C5) were generated using unsupervised hierarchical clustering from the OTU correlation plot. (D) Sample-wise OTU density plot for each cluster (C1, C2, C3, C4, and C5) showing relative abundance. (E-F) CCA plot of microbial community composition for each cluster and bar plot representing cumulative variation percentage from two components [C1 (92.4%), C2 (80.5%), C3 (12.9%), C4 (94.8%), and C5 (39.4%)]. Dotted line shows 30% variance cut-off for downstream analysis.

### Cluster-specific OTUs at genus level identified unique sample-specific OTUs

To gain insight regarding how the bacterial genera were segmented among three groups of samples, we performed genus level OTU correlation (n = 240) and calculated the correlation coefficient (Spearman correlation), followed by unsupervised hierarchical clustering (Figure 3C). We identified 5 distinct clusters, C1 (n = 23), C2 (n = 109), C3 (n = 59), C4 (n = 33) and C5 (n = 16), with variable number of OTUs (Supplemental Table 2). The heat map corresponding to each cluster is shown in Supplemental Figure 4. Cluster-wise OTUs relative abundance density maps were constructed, which distinguished OTUs that were enriched in IA/IS (C1, C3, C4, and C5) and in control samples (C2) (Figure 3D). Some of the enriched cluster-specific OTUs in IA/IS are *Mycobacterium* (1763), *Mycolicibacterium* (1766), *Mycobacteroides* (1774), *Halothiobacillus* (927), *Flavobacterium* (986), *Bifidobacterium* (1695), *Streptomyces* (1884), *Rothia* (2047), and *Mycoplasma* (2100). C2, a control-specific cluster, contains OTUs such as *Thermomicrobium* (500), *Kingella* (502), *Enterobacter* (547), *Bacteroides* (821), and *Prevotella* (840). Thus, this analysis shows the distinction in genus-specific OTUs for both SARS-CoV-2-infected and control subjects. Next, we performed CCA on each of the clusters (C1 to C5) to eliminate sample heterogeneity and enhance the stringency of our analysis pipeline (Figure 3E). We considered the first two components of CCA that explained cumulative variance for the clusters. Cluster C4 explained the highest cumulative variance of 94.8%, while cluster C3 showed the lowest variance of 12.9% (Figure 3F, Supplemental Figure 5A-E). CCA showed efficient sample clustering, which is reminiscent of the density plot (Figure 3D-E).

**Figure 4:**
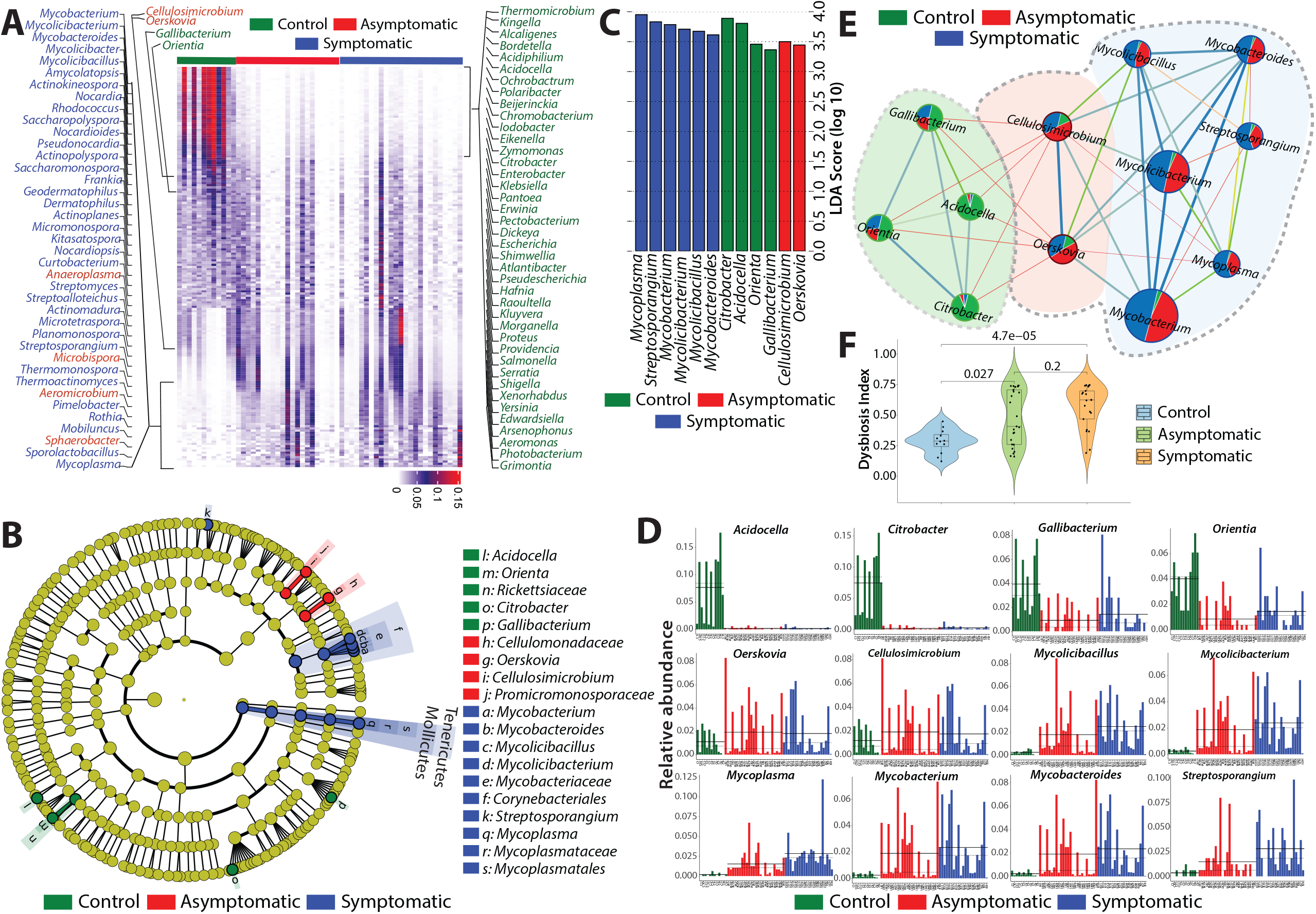
Linear discriminant analysis effect size (LefSe) analysis reveals distinct genus-level OTUs in control, asymptomatic and symptomatic. (A) Heat map showing genus level OTU (n = 181) abundance distribution from four clusters (C1, C2, C4, and C5) identified from CCA analysis in control, asymptomatic and symptomatic samples. The OTUs marked on either side of the heat map were obtained from one-against-all and all-against-all comparison in LDA analysis (B) The cladogram shows the output of the LEfSe (LDA score >2.0), which identifies taxonomic differences between sample groups. Each circle represents a bacterial taxon, and each ring of taxonomy level starting with kingdom in the innermost circle is followed by phylum, class, order, family, and genus in the outermost circle. The different color intensities indicate the different taxonomy levels and the diameter of each circle is proportional to the taxon’s abundance and correlates with the LDA score. (C) The histogram of the LDA scores (score >2.0 and all-against-all) was computed for differentially abundant taxa between sample groups. The effect size of specific taxa in the particular group at the genus level. (D) Histogram of the all LefSe-specific taxa (*Mycoplasma, Streptosporangium, Citrobacter, Acidocella, Mycolicibacterium, Mycolicibacillus, Mycobacterium, Mycobacteroides, Orientia, Gallibacterium, Cellulosimicrobium*, and *Oerskovia*) showing relative abundance across sample groups. Solid and dotted lines show median and mean relative abundance respectively. (E) Weighted correlation network analysis (WGCNA) was used for network construction and plotted using Gephi. Each node of the network represents the individual bacterial genera with their respective abundance size and the edges represent correlation strength with edge weight by thickness. The pie chart within each node represents abundance for each genus. The dotted line shows two distinct modules (control and infected) created in the network analysis. (F) Violin plot showing the dysbiosis indexes of LefSe sample groups (pairwise Wilcoxon rank-sum test p-value < 0.05).

**Figure 5:**
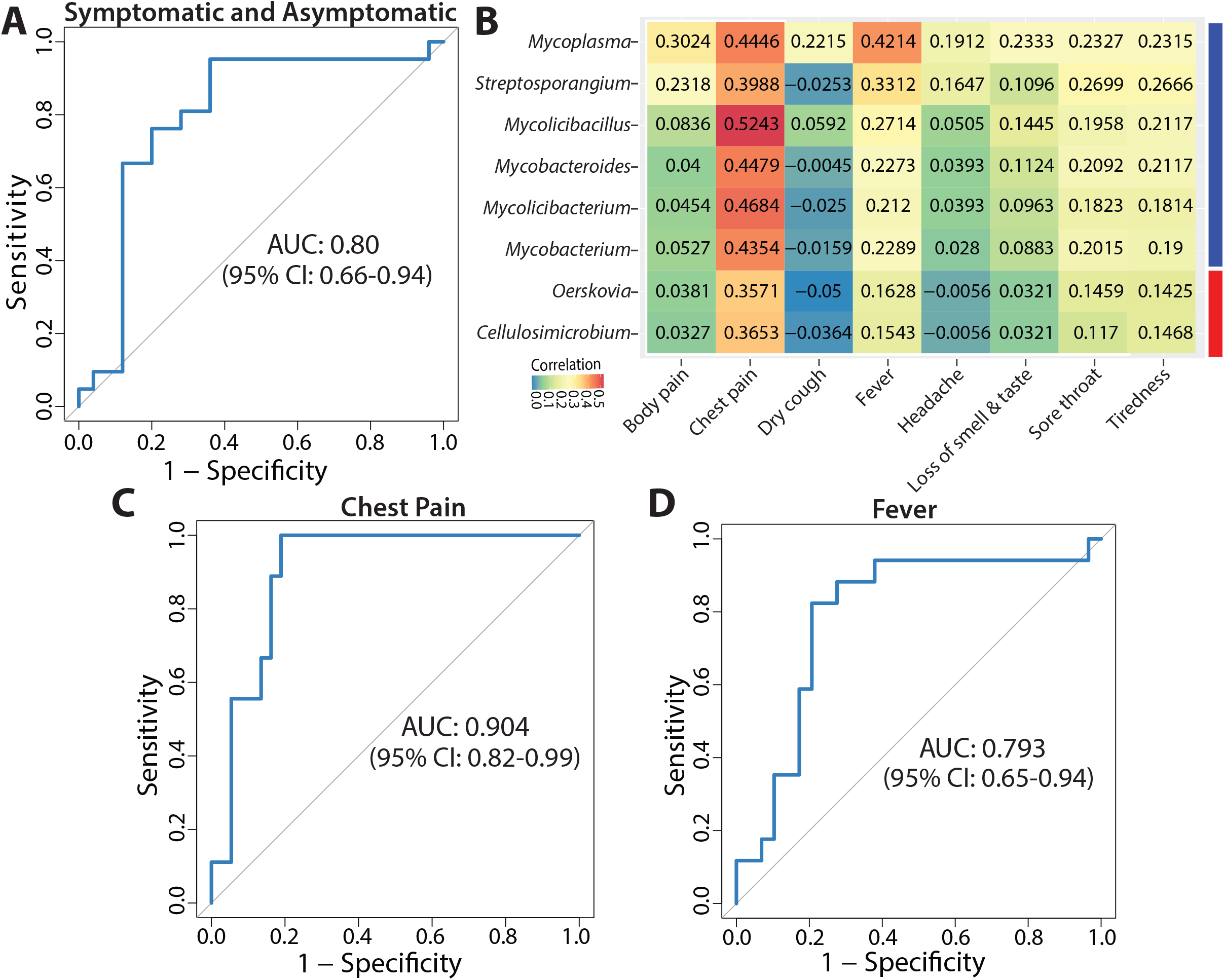
Area under the curve-receiver operating characteristic (AUC-ROC) validation and correlation of genera with the symptoms of COVID-19 subjects. (A) ROC curve for LDA classified symptomatic and asymptomatic group. The AUC w 0.80 with a 95% confidence interval (CI). (B) Correlation between bacteria at genus level and clinical symptoms of patients. (C-D) ROC curve for chest pain and fever in the symptomatic and asymptomatic group. The AUCs were 0.904 (chest pain) and 0.793 (fever) with a 95% confidence interval (CI).

### LefSe analysis identified unique OTUs at genus level in COVID-19 patients

The CCA analysis prompted us to select clusters with maximum variance explained. Therefore, we considered all the clusters with ≥ 30% variance, which includes all the clusters except C3, for LDA. OTUs (n = 181) were extracted from clusters (C1, C2, C4, C5) and plotted in a heat map with their abundance (Figure 4A). Different genera could be clearly distinguished between C, IA, and IS sample groups. Next, we performed LefSe to distinguish the most significant microbiomes from C, IA and IS groups. In a one-against-all comparison (C with IA and IS), we got 40 genera in a control group, 34 genera in the symptomatic group, and 4 genera in an asymptomatic group (LDA score [log10] > 3.6). The genera obtained from one-against-all are highlighted in heatmap (Figure 4A and Supplemental Figure 6A). Relative abundance of each OTU obtained from C, IA, and IS groups are shown in stack plots with clear segregation in OTUs for individual samples (Supplemental Figure 6B-D). The DI calculated from these genera showed high dysbiosis between control and SARS-CoV-2-infected patients (Supplemental Figure 6E-F). We further increased the LefSe stringency by using all-against-all (each sample group compared with each other) comparisons and constructed a cladogram and a bar plot (Figure 4B-C). All the genera obtained from LefSe (One against all and all against all) with their LDA scores and comparison are listed in Supplemental Table 3 and Table 6. We obtained 12 significantly enriched genera of *Gallibacterium, Orientia, Acidocella*, and *Citrobacter* in control samples (LDA score [log10] > 2.0), *Mycoplasma, Streptosporangium, Mycobacterium, Mycolicibacterium, Mycolicibacillus*, and *Mycobacteroides* in symptomatic samples, and *Oerskovia* and *Cellulosimicrobium* in asymptomatic samples (Figure 4C). The histogram showing the relative abundance of the 12 genera for each C, IA, and IS sample group clearly distinguishes each sample type (Figure 4D). Finally, we used weighted correlation network analysis to construct a network (Spearman correlation) with 12 genera identified using the LDA analysis. The network creates two distinct modules, one for control groups and another for both symptomatic and asymptomatic groups. We obtained strong correlation within the genera of C, IA, and IS sample groups (Table 7). However, the correlation between C vs. IA was extremely weak and correlation was not obtained for C vs. IS groups. The network analysis suggested that the NP microbiota of the control group was clearly distinct from that of the asymptomatic and symptomatic groups. The DI of the 12 genera showed the highest significance between C vs. IS (p = 4.7E-05), while significant dysbiosis was not observed between IA and IS groups (Figure 4F). Overall, our analysis confirms the significance of the genera identified and their associations with symptomatic and asymptomatic COVID-19 patients.

**Table 6:**
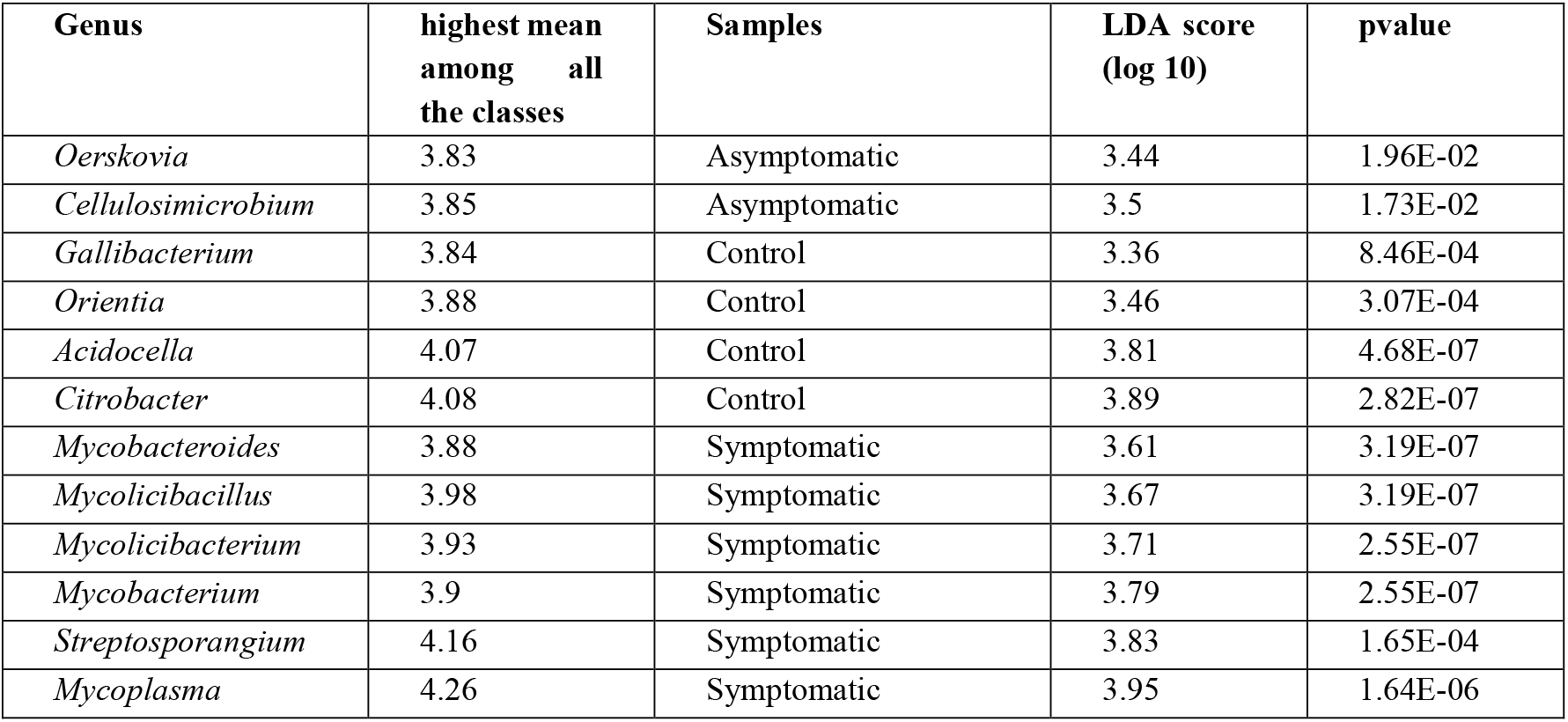
Linear discriminate analysis (LDA) score for all-against-all analysis.

| **Genus** | **highest mean among all the classes** | **Samples** | **LDA score (log 10)** | **pvalue** |
| --- | --- | --- | --- | --- |
| *Oerskovia* | 3.83 | Asymptomatic | 3.44 | 1.96E-02 |
| *Cellulosimicrobium* | 3.85 | Asymptomatic | 3.5 | 1.73E-02 |
| *Gallibacterium* | 3.84 | Control | 3.36 | 8.46E-04 |
| *Orientia* | 3.88 | Control | 3.46 | 3.07E-04 |
| *Acidocella* | 4.07 | Control | 3.81 | 4.68E-07 |
| *Citrobacter* | 4.08 | Control | 3.89 | 2.82E-07 |
| *Mycobacteroides* | 3.88 | Symptomatic | 3.61 | 3.19E-07 |
| *Mycolicibacillus* | 3.98 | Symptomatic | 3.67 | 3.19E-07 |
| *Mycolicibacterium* | 3.93 | Symptomatic | 3.71 | 2.55E-07 |
| *Mycobacterium* | 3.9 | Symptomatic | 3.79 | 2.55E-07 |
| *Streptosporangium* | 4.16 | Symptomatic | 3.83 | 1.65E-04 |
| *Mycoplasma* | 4.26 | Symptomatic | 3.95 | 1.64E-06 |

**Table 7:** WGCNA network data table.

| ID | Genus | OTUs | Module | absolute abundance | Degree | Page ranks | Eigen centrality | Modularity class | Clustering | Triangles |
| --- | --- | --- | --- | --- | --- | --- | --- | --- | --- | --- |
| 1 | *Acidocella* | 525 | M1 | 1362 | 5 | 0.0673 | 0.4963 | 0 | 1 | 10 |
| 2 | *Citrobacter* | 546 | M1 | 3463 | 5 | 0.0673 | 0.4963 | 0 | 1 | 10 |
| 3 | *Gallibacterium* | 750 | M1 | 285 | 5 | 0.0673 | 0.4963 | 0 | 1 | 10 |
| 4 | *Orientia* | 784 | M1 | 263 | 5 | 0.0673 | 0.4963 | 0 | 1 | 10 |
| 5 | *Cellulosimicrobium* | 1710 | M2 | 11832 | 10 | 0.1202 | 1 | 1 | 0.5555 | 25 |
| 6 | *Oerskovia* | 1713 | M2 | 13210 | 10 | 0.1202 | 1 | 1 | 0.5555 | 25 |
| 7 | *Mycobacterium* | 1763 | M2 | 139017 | 7 | 0.0852 | 0.8208 | 1 | 0.9047 | 19 |
| 8 | *Mycolicibacterium* | 1766 | M2 | 81430 | 7 | 0.0852 | 0.8208 | 1 | 0.9047 | 19 |
| 9 | *Mycobacteroides* | 1774 | M2 | 7172 | 7 | 0.0852 | 0.8208 | 1 | 0.9047 | 19 |
| 10 | *Mycolicibacillus* | 1798 | M2 | 2749 | 7 | 0.0852 | 0.8208 | 1 | 0.9047 | 19 |
| 11 | *Streptosporangium* | 2002 | M2 | 163 | 5 | 0.0642 | 0.5732 | 1 | 1 | 10 |
| 12 | *Mycoplasma* | 2100 | M2 | 475 | 7 | 0.0852 | 0.8208 | 1 | 0.9047 | 19 |

### Distinct correlation of OTUs with clinical symptoms in COVID-19 patients

To evaluate the accuracy of LDA classification that identified eight bacterial genera in the IA and IS sample group, we tested the ROC (receiver operating characteristics) – AUC (area under the curve) score. We obtained a value of 0.8 with 95% confidence interval for true positive classification, showing 80% sensitivity and specificity of data obtained from LDA analysis (Figure 5A). Next, we used the Spearman correlation matrix to identify the association of symptoms with the genera. Interestingly, chest pain showed high positive correlation with *Mycoplasma, Mycobacterium, Mycolicibacterium, Mycolicibacillus*, and *Mycobacteroides* which were related to IS group, and weak correlation with *Oerskovia* and *Cellulosimicrobium*, which were associated with the IA group. *Mycoplasma*, however, showed a strong correlation with both chest pain (0.4446) and fever (0.4214) (Figure 5B). ROC-AUC analysis for chest pain and fever showed 0.90 and 0.79 scores, respectively, with eight bacterial genera (Figure 5C-D). We extended our study at the species level for the 12 genera found in LDA analysis and observed that 54 species were represented in a heat map for C, IA, and IS group (Supplemental Figure 7A). Several known opportunistic pathogens such as *Mycobacterium tuberculosis, Mycobacterium avium*, and *Mycoplasma pneumonia* were highly abundant in the SARS-CoV-2-infected patients. The significance of the 54 bacterial species was assessed using the Kruskal-Wallis test and the top 30 significant species were plotted in the bubble plot (Supplemental Figure 7B). In sum, we established the association of pathogenic microbes with COVID-19 disease and showed susceptibility to alterations in the NP microbiome in case of infection in SARS-COV-2. We also identified the compositional difference in NP microbiota between symptomatic and asymptomatic group.

## Discussion

Scientists worldwide are trying to understand the pathophysiology of SARS-CoV-2 infection and the associated alterations in the host, including those in the microbiome. As SARS-CoV-2 infection initiates in the upper respiratory tract, we investigated the alterations in the NP microbiota of COVID-19 patients. We amplified the 16rRNA gene of variable regions (V1-V9) and performed long-read sequencing using Oxford Nanopore technology. Subsequently, we have used multiple bioinformatics approaches to cross-validate our data sets at various levels and identify the most significant bacterial population in the NP microbiome of COVID-19 patients. We found significant changes in abundance, diversity, and DI of SARS-CoV-2-infected patients compared to those of the control. The IA and IS groups also showed overall significant alterations in microbiota composition. We found abundance of opportunistic pathogens such as *Mycoplasma* and *Mycobacterium* in symptomatic patients, which correlated strongly with patient symptoms such as chest pain and fever. Insights into species level abundance revealed the presence of *Mycoplasma pneumoniae, Mycobacterium tuberculosis, Mycobacterium avium*, and *Mycolicibacterium sp*. in the SARS-CoV-2-infected patients. To the best of our knowledge, this is the first comprehensive study to report abundance of opportunistic pathogens such as *Mycoplasma pneumoniae* and *Mycobacterium tuberculosis* based on the complete sequence of the 16S rRNA variable regions in patients with SARS-CoV-2 infection.

Respiratory infections alter the NP microbiota, which reduces the diversity of the NP microbial ecosystem and promotes the growth of opportunistic pathogens (24). At the phylum level, Proteobacteria, Firmicutes, and Actinobacteria were detected in all NP samples. However, the abundance of Firmicutes and Actinobacteria was significantly higher in both symptomatic and asymptomatic groups. Our results are in partial agreement with those of Ventero et al., who found the abundance of Firmicutes, Bacteroidota, Proteobacteria, and Actinobacteria in the NP samples of COVID-19 patients (13). Only few studies have shown either no alterations or significant changes in the microbiome composition of the nasopharynx during COVID-19 infection. Maio et al. and Braun et al. did not find any significant alterations in NP microbial composition (12, 25). However, other studies showed prevalence of opportunistic pathogens such as *Staphylococcus, Anelloviridae, Pseudomonas, Haemophilus, Stenotrophomonas, Redondoviridae*, and *Pseudomonas aeruginosa* in COVID-19 patients (11, 13, 15-18). Compared to earlier reports, our study also revealed overall changes in the composition of the NP microbial community, reduction in bacterial diversity due to COVID-19 infection and the presence of opportunistic pathogens such as *Mycoplasma* and *Mycobacterium* in COVID-19 patient cohort.

Most of the NP microbial studies amplify short 16S rRNA gene using the Illumina platform, which is more accurate but is limited by taxonomic resolution owing to sequencing of shorter reads (26) and sequencing of the specific variable region. The taxonomic resolution can be improved to genus, species, and even at the strain level by sequencing the V1−V9 (∼ 1600 bp) variable regions of the 16S rRNA gene (26, 27). In this study, we have used the Oxford Nanopore™ long read sequenced platform and sequenced V1 to V9 (∼1.6 kb) of 16S variable regions and successfully obtained taxonomic resolution to genus and some extent species level. This has provided us immense advantage of determining the abundance of opportunistic pathogens in the NP of the COVID-19 patients. Until now, only Mostafa et al. has used metagenomics for COVID-19 NP samples using Oxford Nanopore technology. They have sequenced both RNA and DNA from the NP samples without any PCR amplification. They not only identified the SARS-CoV-2 virus in the samples but also potential pathogens that may lead to co-infections (18). Our study is the first to use 16S amplification of ∼1.6 Kb variable regions to identify the bacterial community associated with infected and control NP. However, 16S rRNA gene amplification may introduce PCR biases, however, more subjects and a robust analysis pipeline may dilute these biases. This study is the first comprehensive study from Odisha cohort and second from India. Gupta et al. used the Illumina platform for 16S amplicon sequencing and found enrichment of several opportunistic pathogens (17). Interestingly, *Mycoplasma, Mycolicocibacterium*, and *Mycobacterium* were not present in their list. This could be due to the analysis pipeline or region-specific differences. Nevertheless, the identification of opportunistic pathogens and their increase in abundance in COVID-19 patients is one of the important aspects of this study.

Our comprehensive bioinformatics analysis with sample and OTU correlation analysis distinguished COVID-19 infected and control samples at the genera level. Furthermore, LDA analysis identified a significantly high abundance of *Mycobacterium* and *Mycoplasma* in symptomatic patients, which correlated well with the occurrence of fever and chest pain. Significantly high relative abundance of members of family *Mycobacteriaceae* in the symptomatic COVID-19 group indicates the presence of both pathogenic and non-pathogenic bacteria. Further dissection into genus level revealed the presence of several key genera, namely, *Mycobacterium, Mycolicibacterium, Mycolicibacillus*, and *Mycobacteroides. Mycobacterium* genera are well associated with several pulmonary diseases, for example, *Mycobacterium tuberculosis* is responsible for tuberculosis in humans and is associated with pulmonary infection (28), while *Mycobacterium avium* is highly associated with lung disease (29). *Mycolicibacterium* and *Mycolicibacillus* are generally non-pathogenic but some species have been associated with pathogenicity in humans and were isolated from hospitalized patients (30). *Mycobacteroides* are potentially associated with soft tissue infections and *Mycobacteroides abscessus* is a known pulmonary pathogen (30-32). Members of genus *Mycoplasma* is a well-recognized pathogen, *Mycoplasma pneumoniae* being responsible for pneumonia and other respiratory infections in humans (33, 34). *Cellulosimicrobium* and *Oerskovia* were detected in asymptomatic COVID-19 patients. Few species of *Cellulosimicrobium* are pathogenic, although their pathogenicity was not clear under normal conditions because those species were isolated from hospitalized patients with acute renal failure (35, 36). Results of our and other reports have proved the association of opportunistic pathogens with alterations in the diversity of the microbial communities in symptomatic and asymptomatic COVID-19 patients. This study establishes a new set of opportunistic pathogens in the context of NP microbiome in COVID-19 infected patients. Moreover, this study clearly distinguishes between the NP microbial composition of symptomatic and asymptomatic groups using LefSe with AUC-ROC validation. Thus, we believe that SARS-CoV-2 virulence may promote the growth of opportunistic pathogens and may lead to coinfection or secondary infection in COVID-19 patients.

Our study has certain limitations. The subject size is limited and a larger cohort would have strengthened our findings. The clinical manifestations are limited, and therefore, the larger picture is difficult to interpret. Future studies should include NP samples of vaccinated, asymptomatic, and hospitalized COVID-19 patients with detailed pathophysiology. Furthermore, blood biochemistry and metabolite studies from the serum would boost conclusions regarding functional aspects of the NP microbiome.

## Data Availability

All data produced in the present study are available upon reasonable request to the authors

https://www.ncbi.nlm.nih.gov/sra/PRJNA774098

## Author Contribution

P.P. and A.P. conceptualized the study and secured funding. P.P and S.M. initiated the work, directed overall workflow, interpreted data, and troubleshoot the experiments. R.M. did most of the bioinformatics analysis and S.M., K.C.M. and A.G. helped in bioinformatics analysis and troubleshooting. S.M., S.A., M.S., P.M., R.Y., H.D., S.A.A., S.D., and D.S., helped with the preprocessing of the samples in Biosafety level 3 (BSL3) facility and nucleic acid extractions. S.M., S.A., M.S., P.M., and R.Y. were involved in amplicon library preparations. R.S. provided samples from the Biorepository. A.S., R.D., S.S., T.K.B., S.C., G.H.S., R.S., S.K.R., P.P., and A.P. coordinated with COVID-19 sampling and testing at BSL3. P.P., S.M., and R.M wrote the manuscript.

## Acknowledgments

We acknowledge the institute’s core funding from the Department of Biotechnology (DBT), Government of India. This work was also supported by the ILS flagship project (BT/ILS/Flagship/2019) from DBT, Ramalingaswami Re-entry fellowship (BT/RLF/Re-entry/25/2015), and SERB core research grant (CRG/2018/002052). We also acknowledge Biorepository, BSL-3, and BSL-2 laboratories, qPCR, and DNA-sequencing institutional central core facilities. R.M., S.M., and K.C.M received their fellowships from Ramalingaswami, ILS Flagship, and SERB core research grant, respectively. We thank all the volunteers who provided samples for research purposes.

## References

1. He Y, Wang J, Li F, Shi Y. Main Clinical Features of COVID-19 and Potential Prognostic and Therapeutic Value of the Microbiota in SARS-CoV-2 Infections. Front Microbiol. 2020;11:1302.

2. Khatiwada S, Subedi A. Lung microbiome and coronavirus disease 2019 (COVID-19): Possible link and implications. Hum Microb J. 2020;17:100073.

3. Zou X, Chen K, Zou J, Han P, Hao J, Han Z. Single-cell RNA-seq data analysis on the receptor ACE2 expression reveals the potential risk of different human organs vulnerable to 2019-nCoV infection. Front Med. 2020;14(2):185–92.

4. Hou YJ, Okuda K, Edwards CE, Martinez DR, Asakura T, Dinnon KH, 3rd, et al. SARS-CoV-2 Reverse Genetics Reveals a Variable Infection Gradient in the Respiratory Tract. Cell. 2020;182(2):429–46 e14.

5. Mlcochova P, Kemp SA, Dhar MS, Papa G, Meng B, Ferreira I, et al. SARS-CoV-2 B.1.617.2 Delta variant replication and immune evasion. Nature. 2021.

6. Belkaid Y, Harrison OJ. Homeostatic Immunity and the Microbiota. Immunity. 2017;46(4):562–76.

7. Fazlollahi M, Lee TD, Andrade J, Oguntuyo K, Chun Y, Grishina G, et al. The nasal microbiome in asthma. J Allergy Clin Immunol. 2018;142(3):834–43 e2.

8. de Steenhuijsen Piters WA, Sanders EA, Bogaert D. The role of the local microbial ecosystem in respiratory health and disease. Philos Trans R Soc Lond B Biol Sci. 2015;370(1675).

9. Kumpitsch C, Koskinen K, Schopf V, Moissl-Eichinger C. The microbiome of the upper respiratory tract in health and disease. BMC Biol. 2019;17(1):87.

10. Yildiz S, Mazel-Sanchez B, Kandasamy M, Manicassamy B, Schmolke M. Influenza A virus infection impacts systemic microbiota dynamics and causes quantitative enteric dysbiosis. Microbiome. 2018;6(1):9.

11. Engen PA, Naqib A, Jennings C, Green SJ, Landay A, Keshavarzian A, et al. Nasopharyngeal Microbiota in SARS-CoV-2 Positive and Negative Patients. Biol Proced Online. 2021;23(1):10.

12. De Maio F, Posteraro B, Ponziani FR, Cattani P, Gasbarrini A, Sanguinetti M. Nasopharyngeal Microbiota Profiling of SARS-CoV-2 Infected Patients. Biol Proced Online. 2020;22:18.

13. Ventero MP, Cuadrat RRC, Vidal I, Andrade BGN, Molina-Pardines C, Haro-Moreno JM, et al. Nasopharyngeal Microbial Communities of Patients Infected With SARS-CoV-2 That Developed COVID-19. Front Microbiol. 2021;12:637430.

14. Feehan AK, Rose R, Nolan DJ, Spitz AM, Graubics K, Colwell RR, et al. Nasopharyngeal Microbiome Community Composition and Structure Is Associated with Severity of COVID-19 Disease and Breathing Treatment. Applied Microbiology. 2021;1(2):177–88.

15. Rhoades NS, Pinski AN, Monsibais AN, Jankeel A, Doratt BM, Cinco IR, et al. Acute SARS-CoV-2 infection is associated with an increased abundance of bacterial pathogens, including Pseudomonas aeruginosa in the nose. Cell Rep. 2021;36(9):109637.

16. Nardelli C, Gentile I, Setaro M, Di Domenico C, Pinchera B, Buonomo AR, et al. Nasopharyngeal Microbiome Signature in COVID-19 Positive Patients: Can We Definitively Get a Role to Fusobacterium periodonticum? Front Cell Infect Microbiol. 2021;11:625581.

17. Gupta A, Karyakarte R, Joshi S, Das R, Jani K, Shouche Y, et al. Nasopharyngeal microbiome reveals the prevalence of opportunistic pathogens in SARS-CoV-2 infected individuals and their association with host types. Microbes Infect. 2021:104880.

18. Mostafa HH, Fissel JA, Fanelli B, Bergman Y, Gniazdowski V, Dadlani M, et al. Metagenomic Next-Generation Sequencing of Nasopharyngeal Specimens Collected from Confirmed and Suspect COVID-19 Patients. mBio. 2020;11(6):1–13.

19. Liu J, Liu S, Zhang Z, Lee X, Wu W, Huang Z, et al. Association between the nasopharyngeal microbiome and metabolome in patients with COVID-19. Synth Syst Biotechnol. 2021;6(3):135–43.

20. Haiminen N, Utro F, Seabolt E, Parida L. Functional profiling of COVID-19 respiratory tract microbiomes. Sci Rep. 2021;11(1):6433.

21. Wood DE, Lu J, Langmead B. Improved metagenomic analysis with Kraken 2. Genome Biol. 2019;20(1):257.

22. Lloyd-Price J, Arze C, Ananthakrishnan AN, Schirmer M, Avila-Pacheco J, Poon TW, et al. Multi-omics of the gut microbial ecosystem in inflammatory bowel diseases. Nature. 2019;569(7758):655–62.

23. Bastian M, Heymann S, Jacomy M. Gephi: An Open Source Software for Exploring and Manipulating Networks. Proceedings of the International AAAI Conference on Web and Social Media. 2009;3(1):361–2.

24. Santacroce L, Charitos IA, Ballini A, Inchingolo F, Luperto P, De Nitto E, et al. The Human Respiratory System and its Microbiome at a Glimpse. Biology (Basel). 2020;9(10).

25. Braun T, Halevi S, Hadar R, Efroni G, Glick Saar E, Keller N, et al. SARS-CoV-2 does not have a strong effect on the nasopharyngeal microbial composition. Sci Rep. 2021;11(1):8922.

26. Johnson JS, Spakowicz DJ, Hong BY, Petersen LM, Demkowicz P, Chen L, et al. Evaluation of 16S rRNA gene sequencing for species and strain-level microbiome analysis. Nat Commun. 2019;10(1):5029.

27. Kaul D, Rathnasinghe R, Ferres M, Tan GS, Barrera A, Pickett BE, et al. Microbiome disturbance and resilience dynamics of the upper respiratory tract during influenza A virus infection. Nat Commun. 2020;11(1):2537.

28. Peto HM, Pratt RH, Harrington TA, LoBue PA, Armstrong LR. Epidemiology of extrapulmonary tuberculosis in the United States, 1993-2006. Clin Infect Dis. 2009;49(9):1350–7.

29. Hwang JA, Kim S, Jo KW, Shim TS. Natural history of Mycobacterium avium complex lung disease in untreated patients with stable course. Eur Respir J. 2017;49(3):1600537.

30. Gupta RS, Lo B, Son J. Phylogenomics and Comparative Genomic Studies Robustly Support Division of the Genus Mycobacterium into an Emended Genus Mycobacterium and Four Novel Genera. Front Microbiol. 2018;9:67.

31. Batchelder HR, Story-Roller E, Lloyd EP, Kaushik A, Bigelow KM, Maggioncalda EC, et al. Development of a penem antibiotic against Mycobacteroides abscessus. Commun Biol. 2020;3(1):741.

32. Tortoli E. Microbiological features and clinical relevance of new species of the genus Mycobacterium. Clin Microbiol Rev. 2014;27(4):727–52.

33. Beeton ML, Zhang XS, Uldum SA, Bebear C, Dumke R, Gullsby K, et al. Mycoplasma pneumoniae infections, 11 countries in Europe and Israel, 2011 to 2016. Euro Surveill. 2020;25(2).

34. Foy HM. Infections caused by Mycoplasma pneumoniae and possible carrier state in different populations of patients. Clin Infect Dis. 1993;17 Suppl 1:S37–46.

35. Sharma A, Gilbert JA, Lal R. (Meta)genomic insights into the pathogenome of Cellulosimicrobium cellulans. Sci Rep. 2016;6:25527.

36. Delport J, Wakabayashi AT, Anantha RV, Lannigan R, John M, McCormick JK. Cellulosmicrobium cellulans isolated from a patient with acute renal failure. JMM Case Reports. 2014;1(2):e000976.

