## Supplemental data for "Nanopore 16S rRNA sequencing reveals alterations in nasopharyngeal microbiome and enrichment of *Mycobacterium* and *Mycoplasma* in patients with COVID 19"

### <u>Supplementary Appendix</u>

#### Supplemental Figure S1

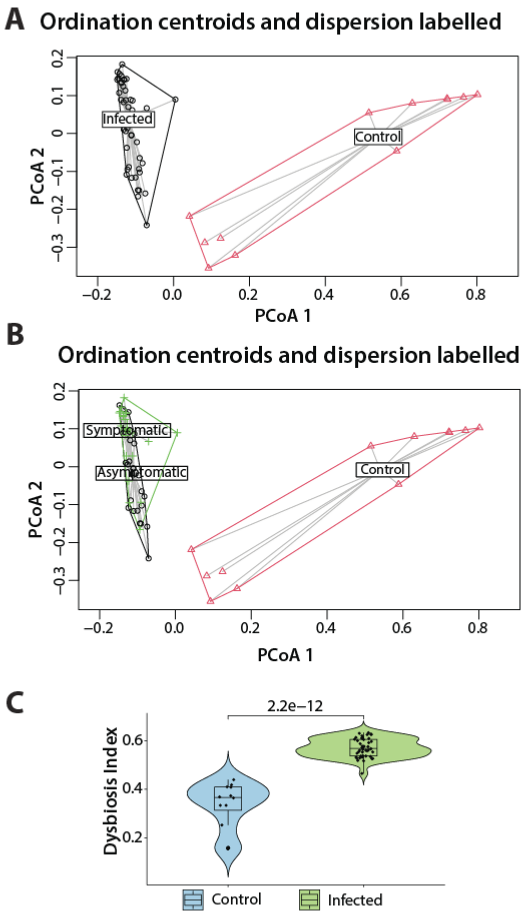

**Figure S1: Centroid and Euclidian distance for dysbiosis index calculation in control and COVID-19 subjects.** (A-B) Principal co-ordinate analysis centroid ordination of sample groups. Dysbiosis index calculated for each sample based on Euclidean distance from the centroid of the control sample. (C) The violin plot shows the dysbiosis indexes of samples from control and infected individuals (Wilcoxon rank sum test p value < 0.05).

#### Supplemental Figure S2

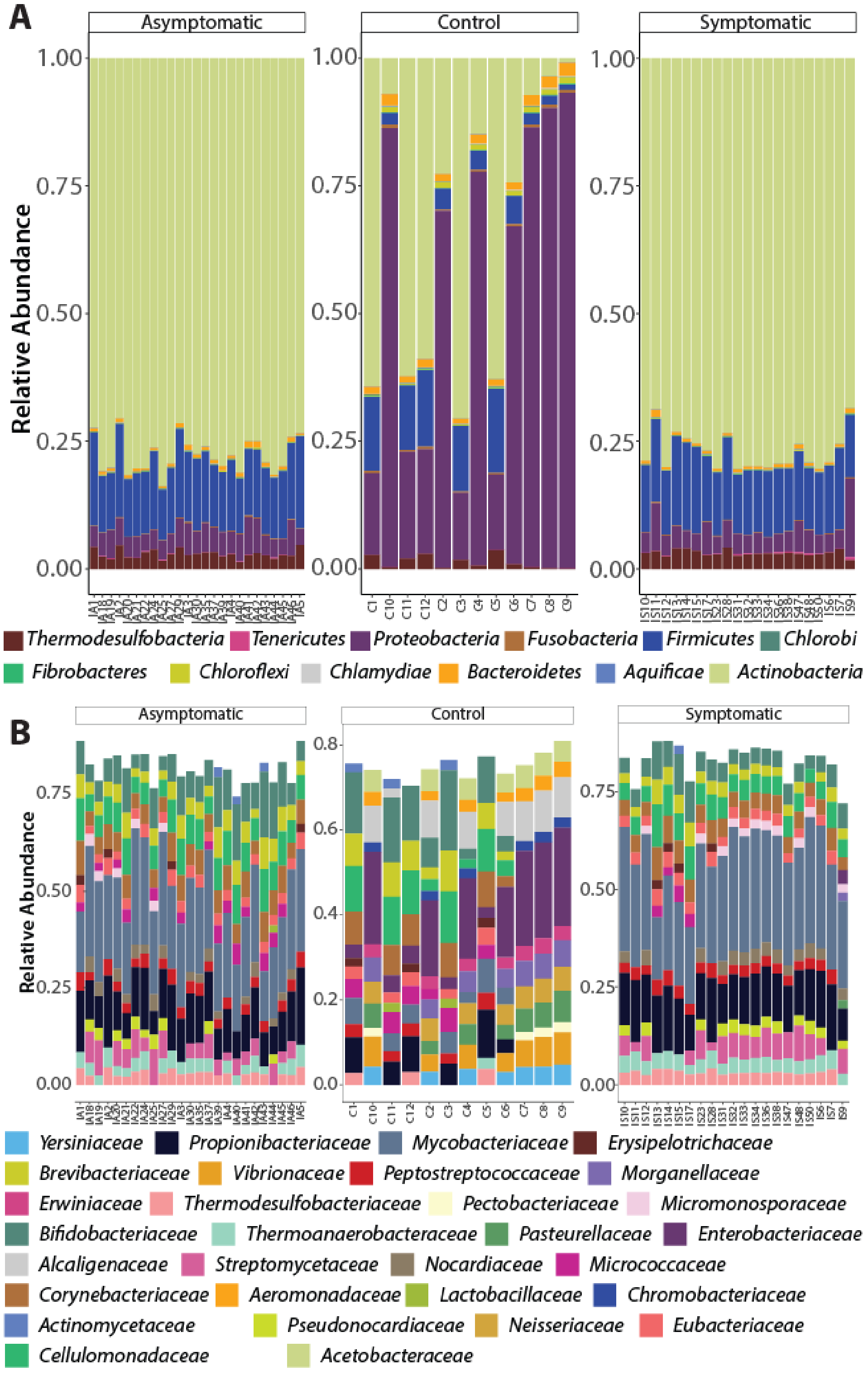

**Figure S2: Relative abundance of major phyla and family between control, asymptomatic, and symptomatic COVID-19 participants**. (A-B) Stack plots showing relative abundance of phylum and family OTUs obtained in control, asymptomatic, and symptomatic COVID-19 participants.

**Supplemental Figure S3**

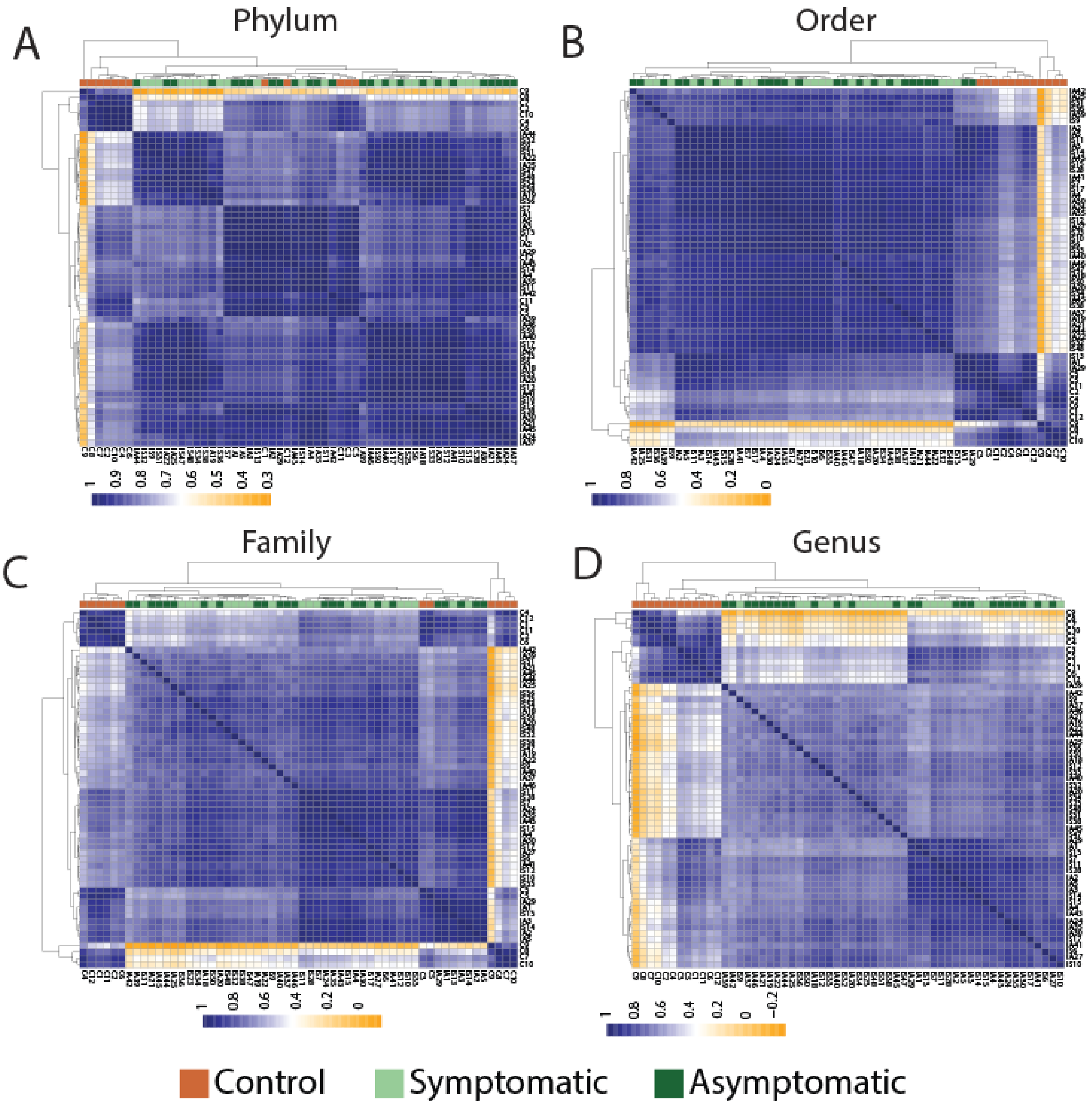

**Figure S3: Correlation plots across taxonomic classification.** (A-D) Heat map of sample-sample correlation using Spearman correlation method for phylum, order, family, and genus. Sample segregation and clustering is shown for control (orange), symptomatic (light green), and asymptomatic (dark green) cases in all plots.

**Supplemental Figure S4**

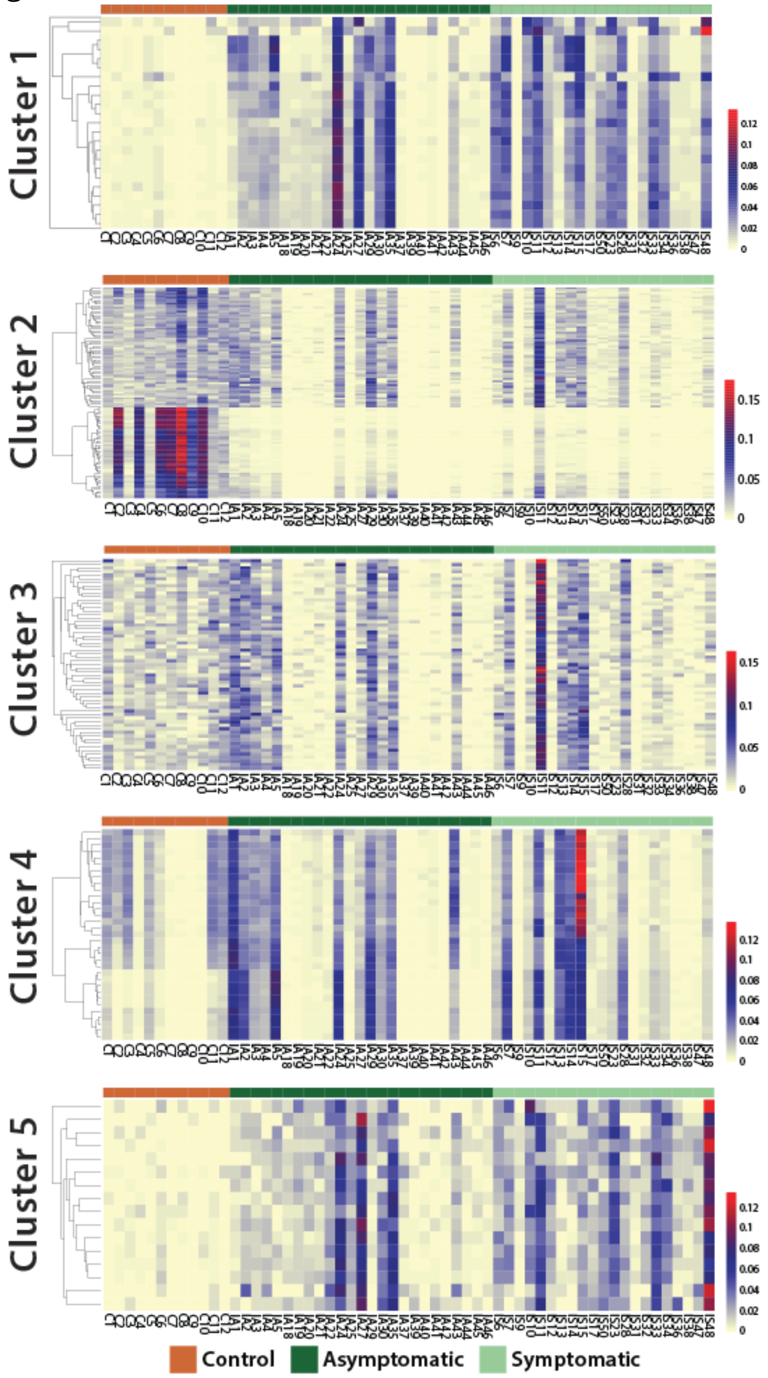

**Figure S4: Cluster-specific heat maps for all five clusters obtained from OTU-OTU correlation analysis.** Heat map showing relative abundance of genus level OTUs across control (orange), asymptomatic (dark green), and symptomatic (light green) participants. The OTU numbers in different clusters are: C1, n = 23; C2, n = 109; C3, n = 59; C4, n = 33; and C5, n = 16.

**Supplemental Figure S5**

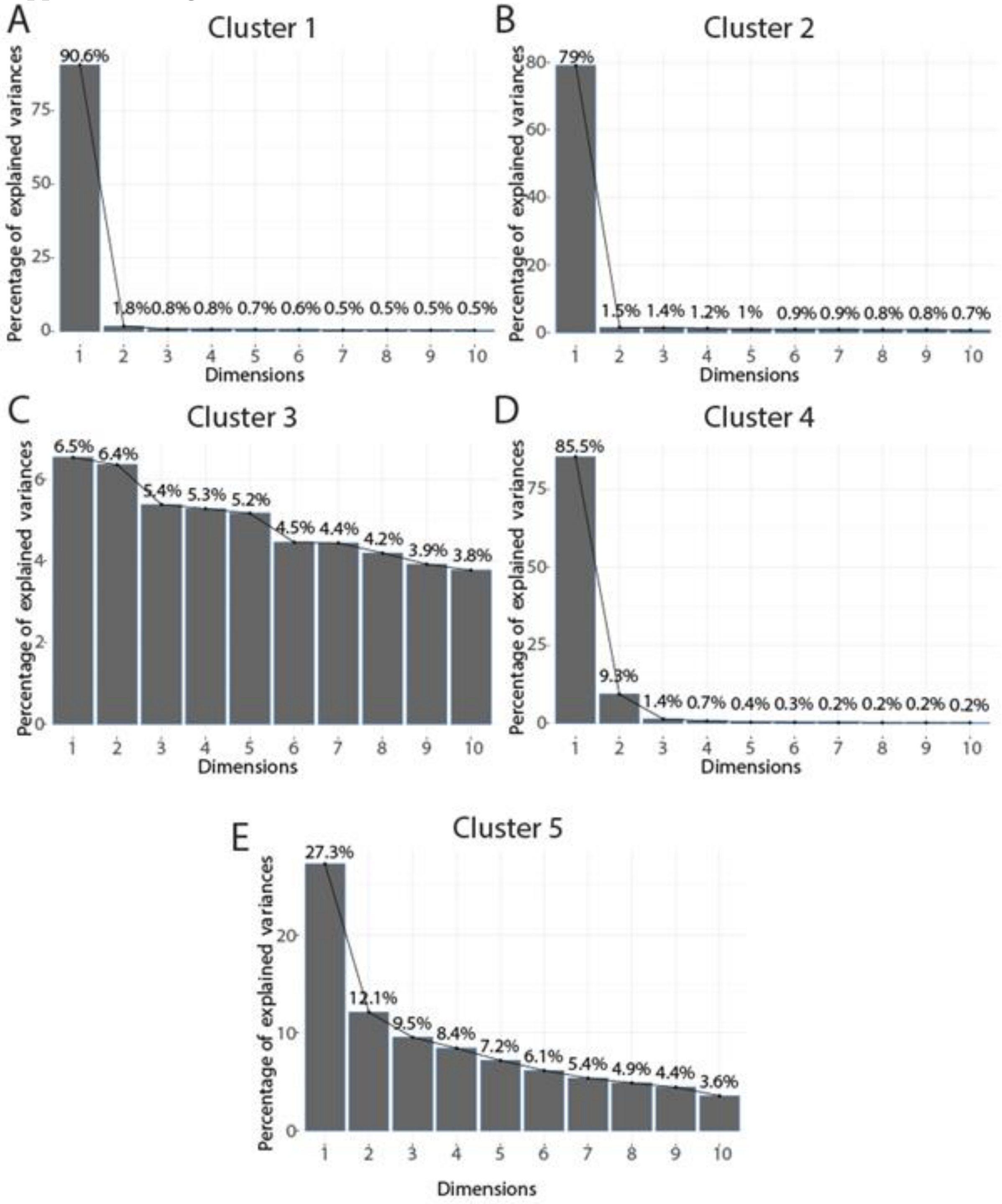

**Figure S5: Scree plots for all five clusters obtained from OTU-OTU correlation analysis.** (A-E) Scree plot of five clusters (C1-C5) from CCA showing the percentage of total variance in the clusters as explained or represented by each component.

#### Supplemental Figure S6

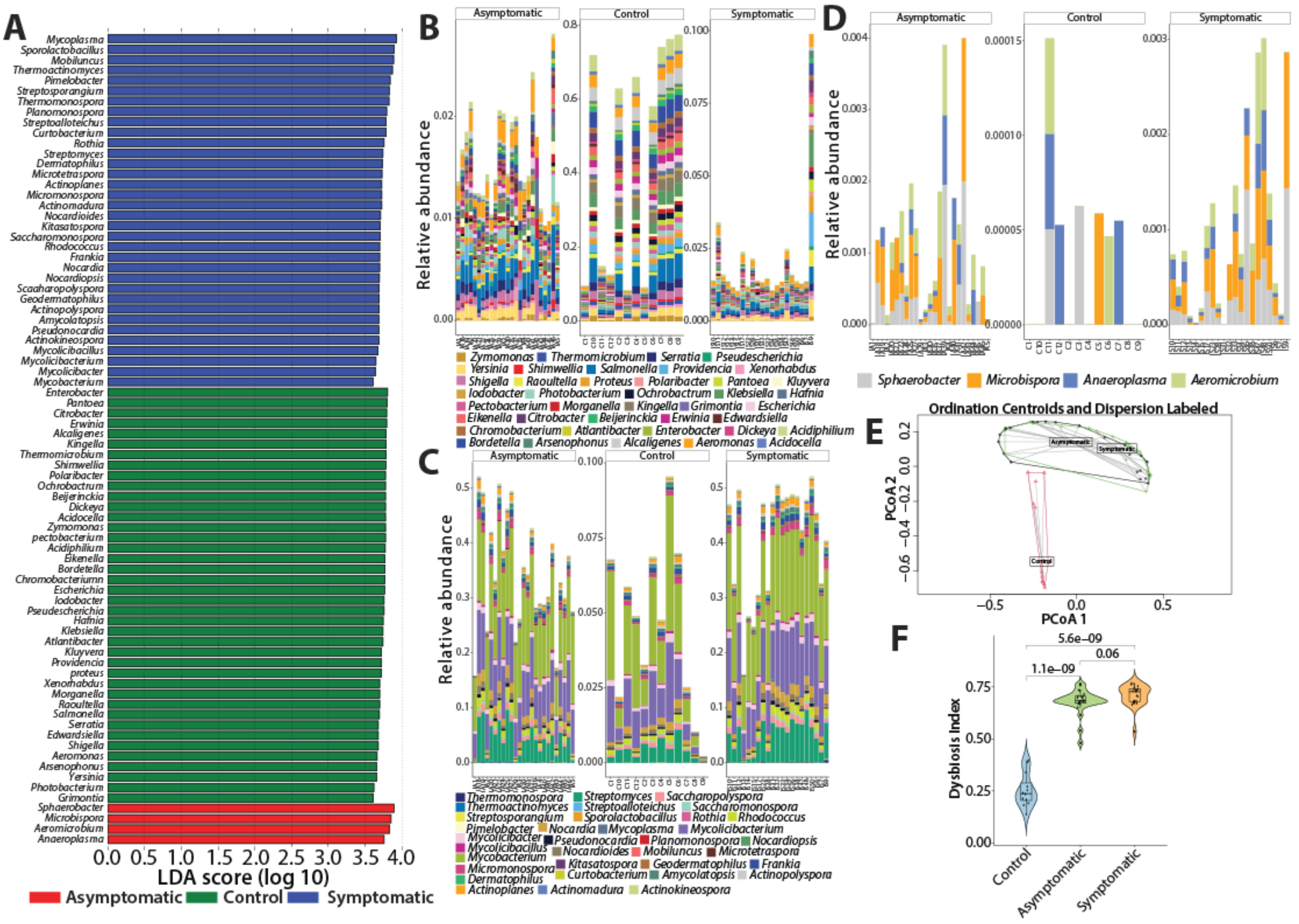

**Figure S6: Linear discriminant analysis effect size (LefSe) analysis comparing control with symptomatic and asymptomatic cases (one-against-all) and dysbiosis index.** (A) Histogram of the LDA Scores (cut-off >3.6) was computed for differentially abundant OTUs at genus level between three sample groups. The bar size represents the effect size of each taxa at genus level for each sample group. (B-D) Stack plots showing OTU abundance obtained from LDA analysis for control, symptomatic, and asymptomatic cases. (E-F) Dysbiosis index calculated from genus-specific OTUs obtained from LDA analysis for control, asymptomatic, and symptomatic samples (pairwise Wilcoxon rank sum test $p \leq 0.05$).

**Supplemental Figure S7**

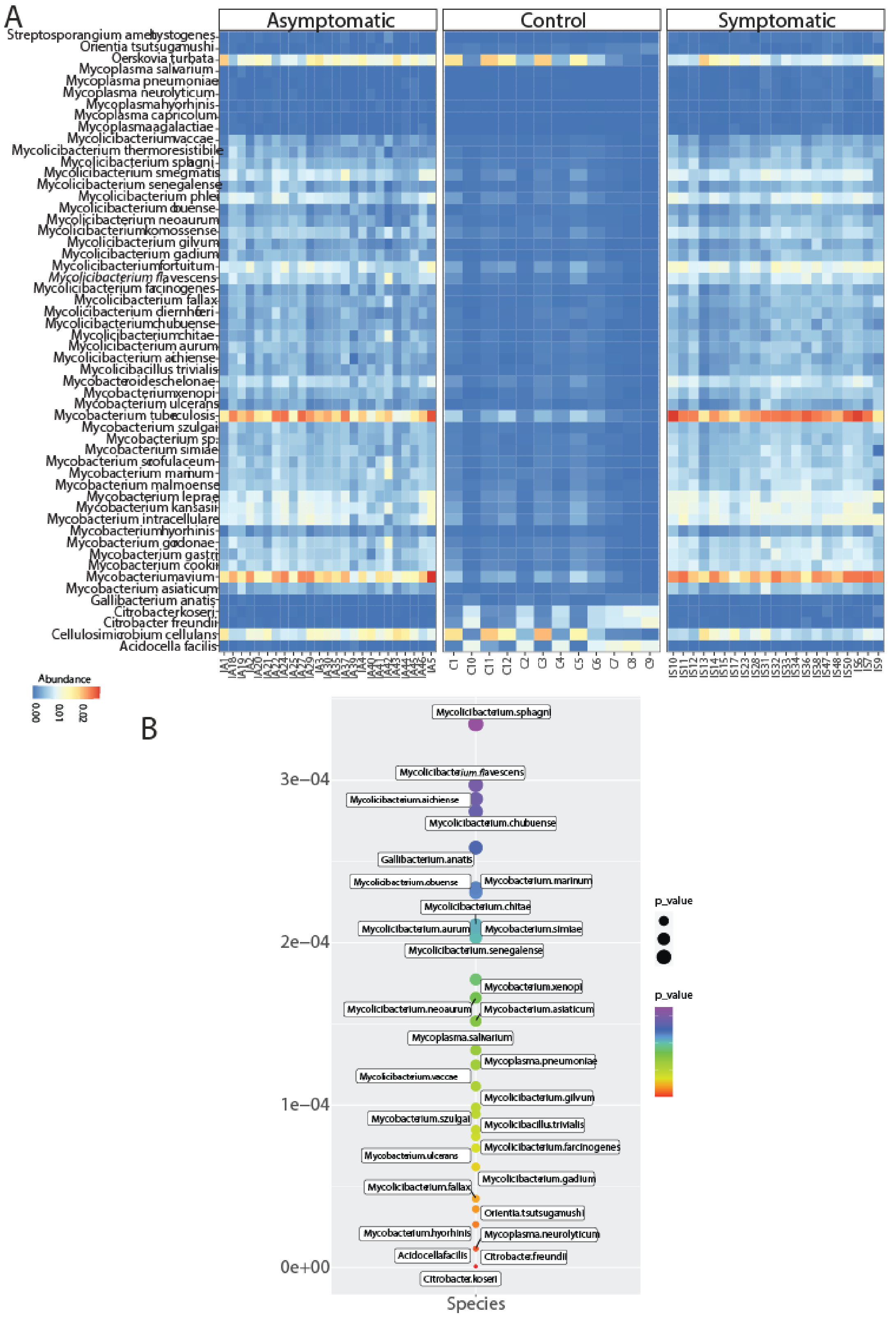

**Figure S7: Relative abundance and significance of species-specific OTUs.** (A) Heat map showing the taxonomic distribution of major species based on LEfSe results within sample groups. (B) Bubble plot showing the Kruskal-Wallis test significance of p-values of each bacterial species.
